# Efficiencies in the allocation of insecticide-treated nets for malaria prevention in urban Sub-Saharan Africa

**DOI:** 10.1101/2025.05.29.25328568

**Authors:** Colleen Leonard, Ifeoma D. Ozodiegwu, Anne Stahlfeld, Chilochibi Chiziba, Nicholas S. Adam, Ousmane Oumou Diallo, Beatriz Galatas, Jaline Gerardin, Abdisalan Noor

## Abstract

The extent to which insecticide-treated nets (ITNs), a key malaria prevention intervention in Sub-Saharan Africa, have been appropriately targeted to those at highest risk remains unknown, particularly within the urban context. We combined household surveys, programmatic ITN distribution data, and procurement cost data to estimate the number and cost of ITNs distributed in urban areas of Sub-Saharan Africa over the period 2010-2021, as well as trends in malaria prevalence and ITN access by wealth quintile. We estimate that over 440 million ITNs at a cost of over $1B USD have been distributed in urban areas, and allocations between rural and urban areas did not proportionately reflect the higher risk of malaria in rural areas. In urban areas, households in lower wealth quintiles were at higher risk of malaria but did not have greater access to ITNs. Housing quality and wealth but not ITN access or use were associated with decreased malaria prevalence in urban areas. If ITNs in urban areas had been distributed solely to slum residents, over $200 million could have been freed for other malaria control activities. In the context of needing to prioritize increasingly limited resources for health, ITN distributions must be better targeted to the most vulnerable.

## Background

Sub-Saharan Africa bears the largest burden of malaria worldwide, accounting for approximately 95% of all cases^1^. In general, malaria transmission is lower in urban areas than in rural areas. Several environmental, socioeconomic, and behavioral factors drive differences in transmission across urban and rural areas^2^. In urban areas, better infrastructure often reduces breeding sites and prevents mosquitoes from coming indoors, and urban residents tend to have greater access to healthcare and treatment for malaria^3-5^. Use of insecticide-treated nets (ITNs) is typically moderate to low in urban areas compared to rural areas due to perceived and actual lower risk^2^. Outdoor biting may also be more common in urban settings^6^. Furthermore, travel from rural areas can drive malaria in urban centers of sub-Saharan Africa^7,8^. Indoor-based vector control strategies such as ITNs may therefore have limited impact on malaria transmission in some urban settings. In the context of limited resources, reallocating funds away from ITN distributions in urban centers toward other malaria needs may result in increased impact.

At the same time, malaria transmission is often heterogeneous within urban areas, with some urban residents at higher risk than others. Rapid urbanization, which is occurring in Africa^9^, can create areas with inadequate housing and poor sanitation. Informal settlements and slums with overcrowded conditions, poor sanitation, and poor housing conditions have been associated with higher malaria transmission compared to other areas of a city^10^. Slum residents may have particularly poor access to ITNs^11^. If populations at highest risk within cities can be identified, this would better inform malaria prevention strategies in urban areas considering limited funds.

While national-level data on number of ITNs distributed is reported annually^12^, there has not yet been any effort to understand the investment on ITNs made in urban sub-Saharan Africa. Here, we use publicly available data to explore the allocation of ITNs in the region’s urban context. Given the limitations of the data available at the continental scale, we focus on back-of-the-envelope approaches that give a general idea of patterns across sub-Saharan Africa. We estimate how many ITNs have been distributed in urban areas of sub-Saharan Africa from 2011-2021, the total funds spent on their procurement, and their distribution by wealth quintile. We characterize trends in malaria prevalence and ITN access in urban settings, distinguishing between different types of urban areas and urban households. Finally, we present counterfactuals to explore how funds for urban ITNs could have instead been used for other interventions over the period 2011-2021.

## Results

### Resources allocated to ITNs in urban and rural areas between 2011 and 2021

Since 2011, malaria prevalence in children under five years of age in rural areas of sub-Saharan Africa has been 3 to 5-fold higher than in urban areas (Figure 1A, Figure S1). This difference persisted even in recent surveys (2016-2021), which reported a median prevalence of 6% in urban areas and 28% in rural areas.

**Figure 1.**
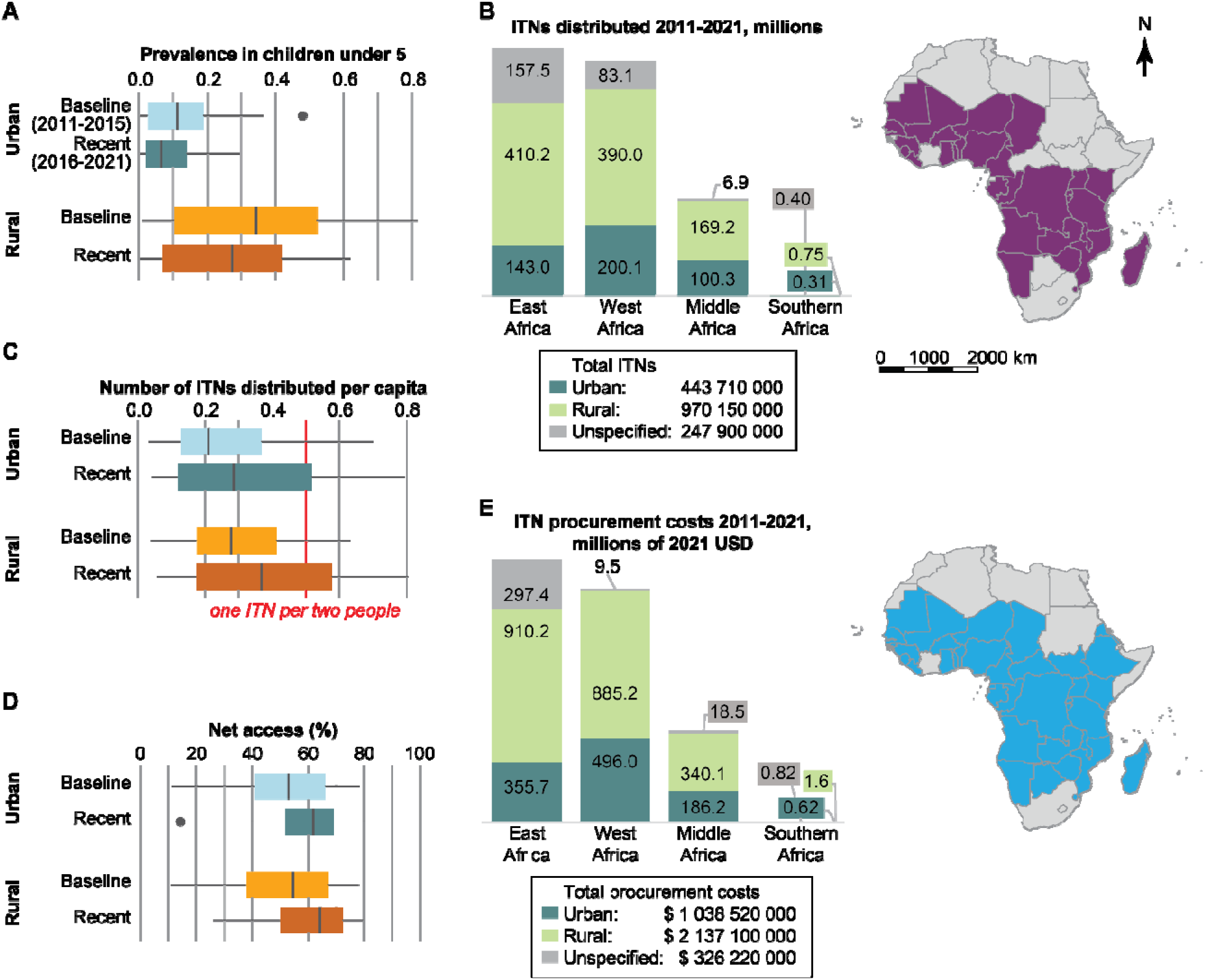
Malaria prevalence and ITN deployment in urban and rural areas of sub-Saharan Africa, 2011-2021, at the continental and regional level. A) Prevalence of malaria infection in children under 5 over the last decade, stratified by residence (urban or rural) and survey period (2011-2015 for baseline, or 2016-2021 for recent), among 23 sub-Saharan African countries with at least one DHS or MIS survey between 2011 and 2021 (Figure S1, Table S1). All boxplots indicate median and interquartile range. The black dots represent outliers defined as 1.5*inter-quartile range. B) Total ITNs distributed in sub-Saharan Africa, 2011-2021, in rural and urban sites stratified by region, and countries included in this estimate. C) Number of ITNs distributed per capita in urban and rural areas, during baseline and recent periods. D) Percent of urban and rural residents with access to an ITN, assuming one ITN per two people, during baseline and recent periods. E) Total estimated costs of ITN procurement in sub-Saharan Africa 2011-2021, stratified by rural and urban areas, and countries (Figure S9) included in this analysis.

Over the period 2011-2021, in 31 countries with ITN ownership data available, over 440 million ITNs were distributed to urban areas, compared to 970 million distributed to rural areas (Figure 1B). Country-specific distribution numbers per year are shown in Figure S2. Per capita, rural residents received slightly more ITNs than urban residents (Figure 1C, Figure S3), with resulting ITN access similar across urban and rural areas (Figure 1D) rather than reflecting the substantial higher risk of malaria in rural areas. ITN access here refers to the proportion of the population that could sleep under an ITN, assuming one ITN per two people. Even in recent years, ITN distribution was insufficient to reach one ITN per two rural residents or achieve universal access.

In 43 countries where ITN costs could be estimated, over $1.0 billion U.S. dollars were spent on ITN procurement in urban areas from 2011-2021, compared with over $2.1 billion dollars spent on ITNs in rural areas (Figure 1E).

### Heterogeneity of malaria risk in urban areas

Between individual countries, malaria prevalence in children under five living in urban areas varied widely, ranging from 0.1% (Senegal, 2014 & 2016) to 47.9% (Burkina Faso, 2010) (Figure 2). From 2010-2021, eleven countries experienced a decrease in urban malaria prevalence (Burkina Faso, Cameroon, Gambia, Ghana, Guinea, Kenya, Malawi, Mali, Nigeria, Tanzania, Uganda), three countries experienced an increase (Angola, Benin, and Burundi), and urban malaria prevalence remained relatively stable in seven countries (Cote d’Ivoire, Liberia, Madagascar, Mozambique, Rwanda, Senegal, and Togo).

**Figure 2.**
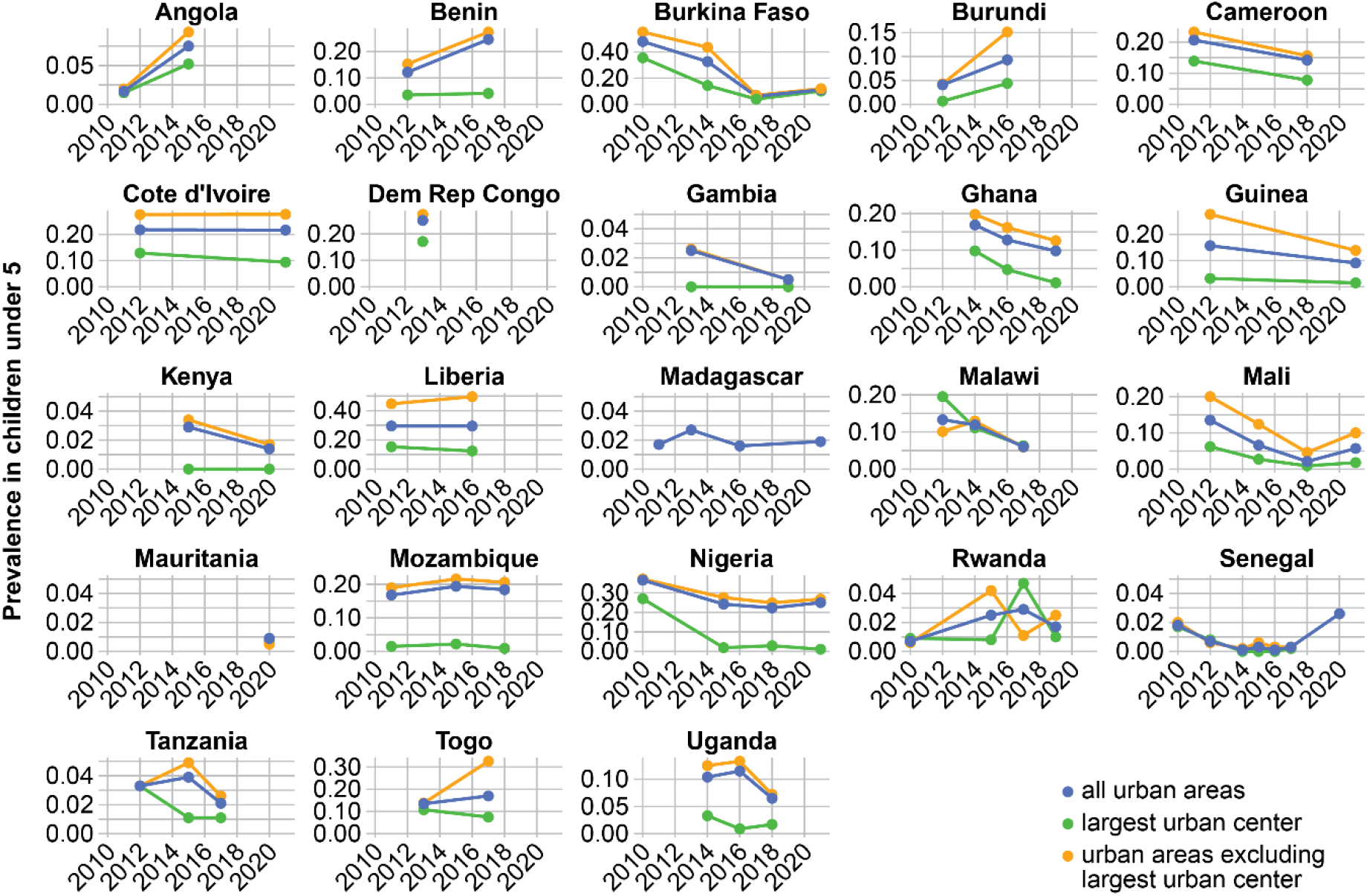
Malaria prevalence in children under 5 by RDT in urban areas, largest urban center, and the remaining urban areas (excluding the largest urban center) by country in sub-Saharan Africa. Points indicate data from each survey.

Malaria prevalence in children under 5 in urban areas was heterogeneous within countries. In all countries and survey years (except Rwanda 2017), the largest urban center had much lower prevalence than other urban areas in the country, occasionally substantially so (for example, Liberia and Mozambique). Trends could also differ, as in Benin where prevalence in the largest urban center remained stable at 4% while prevalence in the remaining urban areas almost doubled from 15 to 27% between 2012 and 2017.

Urban malaria prevalence was always higher among children living in households in the lower three wealth quintiles (except Burundi 2016) compared to those in the highest two wealth quintiles (Figure 3). The gap narrowed among recent surveys for most countries, although all countries except Burkina Faso, Burundi, Gambia, Mauritania, and to some extent Tanzania continued to show substantially higher prevalence amongst children from poorer households.

**Figure 3.**
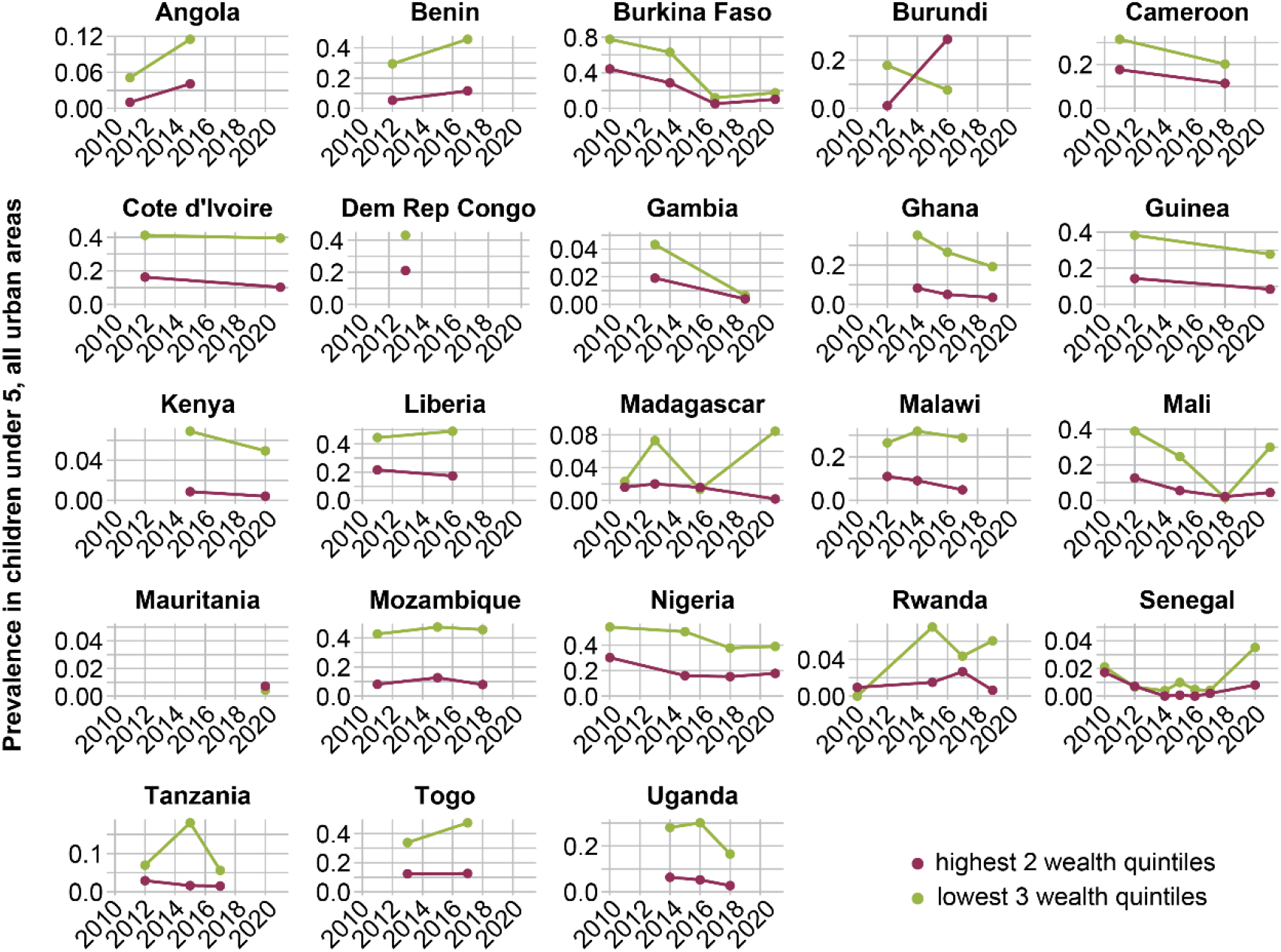
Malaria prevalence in children under 5 by RDT in urban areas, stratified by household wealth quintile, by country. Points indicate data from each survey.

### ITN access in urban areas

ITN access in urban areas, defined as the proportion of the population that could have slept under an ITN the previous night under the assumption of one net covering two household members, varied widely by country from 2010-2021 (Figure 4, Figure S4), ranging from 11% in Cameroon (2011) to 81% in Benin (2017), with mean access of 60% among the most recent surveys between 2016-2021. ITN access and use was often, though not always, higher in urban areas other than the largest urban center (Figure S5). Over the study period, ITN access in urban areas increased in 12 countries (Benin, Burkina Faso, Cameroon, Côte d’Ivoire, Gambia, Guinea, Liberia, Malawi, Mozambique, Nigeria, Senegal, Togo), decreased in four (Burundi, Kenya, Madagascar, Tanzania), and remained stable or had indeterminate trend in the remaining seven.

**Figure 4.**
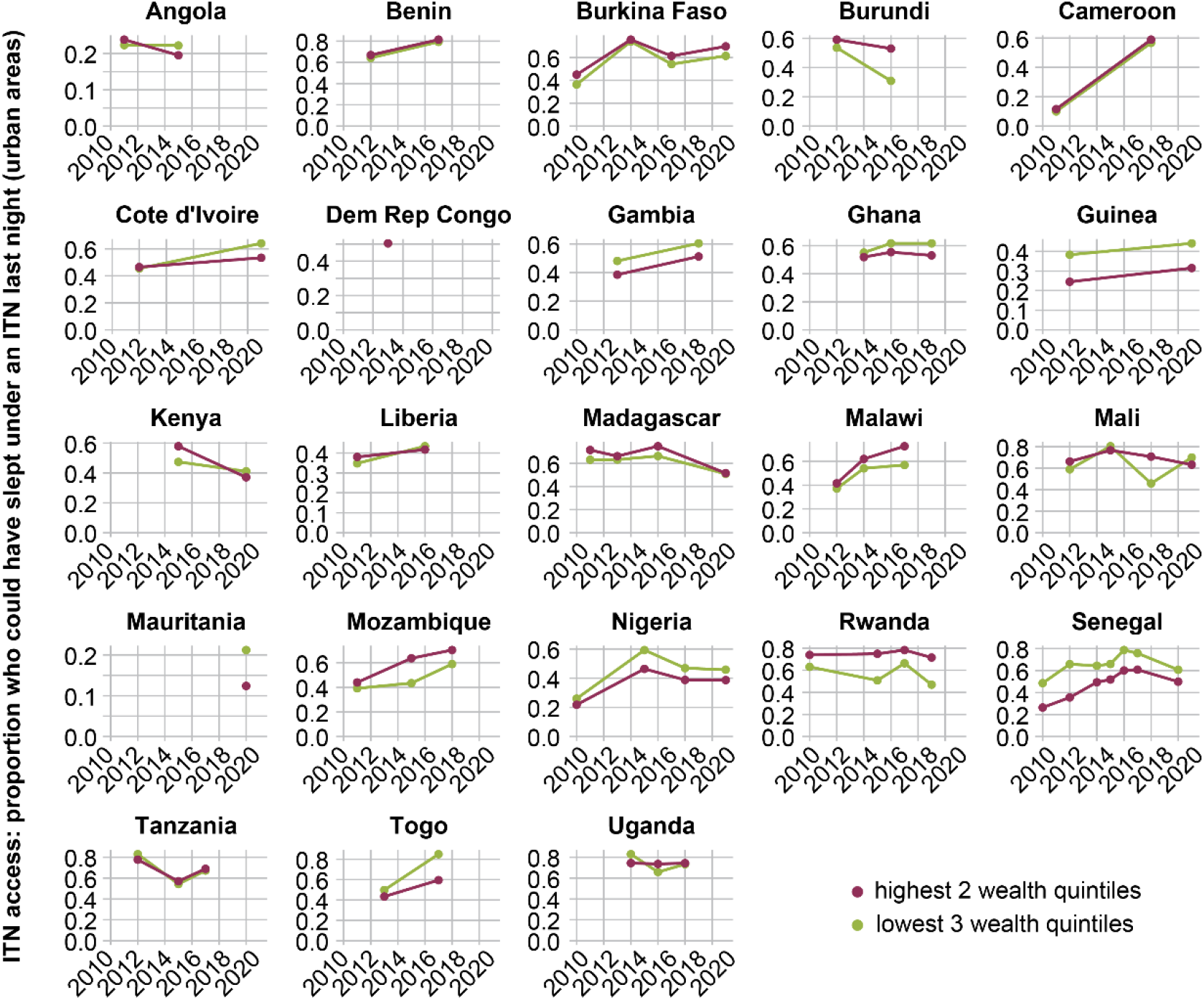
ITN access in urban areas, defined as the proportion of the population that could have slept under an ITN the previous night assuming 1 ITN per 2 household members, stratified by wealth quintile. Points indicate data from each survey.

In six countries (Gambia, Ghana, Guinea, Nigeria, Senegal, Togo), urban residents in lower wealth quintiles had consistently higher access to ITNs than those in higher wealth quintiles, although their increased access to ITNs was generally still not as large as their increased malaria risk (Figure 3). In four countries (Burundi, Malawi, Mozambique, and Rwanda), urban residents in lower wealth quintiles had consistently lower access to ITNs than those in higher wealth quintiles. In the remaining thirteen countries, access to ITNs was similar across household wealth strata.

In Côte d’Ivoire and Togo, access for less wealthy households improved more over time than access for wealthier households, but for all other countries, there is no evidence that poorer residents were better targeted with ITN distributions in recent years. ITN use in urban areas, defined as the proportion of the population that slept under an ITN the night before the survey, stratified by household wealth followed similar patterns as ITN access (Figure S6). In 16 out of 23 countries, urban residents in lower wealth quintiles were more likely to have used a net to which they had access, compared to residents in higher wealth quintiles.

### Factors associated with malaria risk in urban areas

Increases in urban ITN use occasionally coincided with decreases in urban malaria prevalence over the same period (e.g. Cameroon), but not always (e.g. Kenya). To assess how various factors may be associated with urban malaria, we used a zero-inflated Poisson regression model to model malaria test positivity in urban clusters over time. Enhanced vegetation index (EVI) was positively associated with malaria infections in urban areas (Table 1). Both modern housing (RR= 0.70 (95% CL: 0.64, 0.76)) and household wealth (0.38 (95% CL: 0.35, 0.41)) were significantly negatively associated with malaria positivity. Neither ITN access nor ITN use had a significant association. After accounting for the model covariates, predicted urban malaria positivity generally decreased from 2010 to 2021 (Figure S7).

**Table 1.** Zero-inflated Poisson regression estimates for malaria test positivity rate in an urban cluster.

| Variables | Rate ratio | 95% confidence limit |
| --- | --- | --- |
| Net use | 1.09 | (0.91, 1.31) |
| Net access | 0.88 | (0.73, 1.06) |
| Modern housing | 0.70 | (0.64, 0.76)* |
| Household wealth† | 0.38 | (0.35, 0.41)* |
| Monthly total precipitation | 1.06 | (0.87, 1.29) |
| Enhanced vegetation index | 7.77 | (6.47, 9.34)* |
| Relative humidity | 0.99 | (0.99, 1.00) |
| Year | 0.95 | (0.94, 0.96)* |

†Household wealth is defined as wealthy (1) for the highest and second highest wealth quintiles, and not wealthy (0) for the lower three wealth quintiles.
\*Statistically significant at $p < 0.05$ .

### Potential savings from targeting urban ITNs only to slum residents

Given the higher malaria risk in poorer urban households, a targeted ITN distribution strategy in urban areas should prioritize those in the lowest wealth quintile, such as slum residents. However, we found a similar number of ITNs across wealth quintiles in urban areas (Figure 5A), suggesting that poorer populations may not have been meaningfully targeted for distribution over those at lower risk.

**Figure 5.**
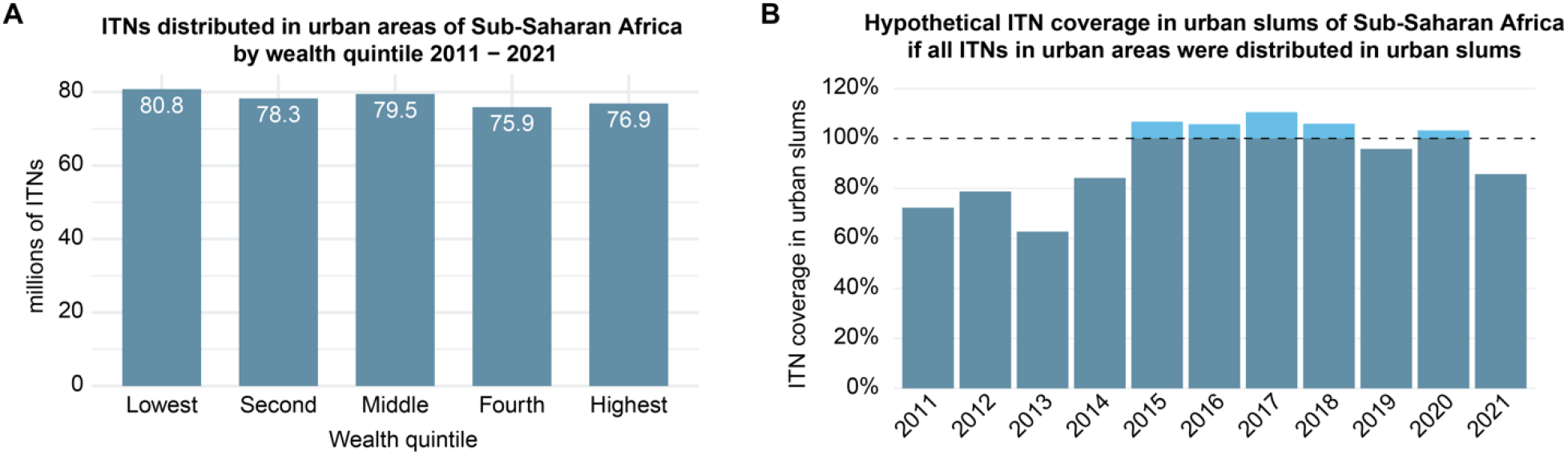
ITN distributions by wealth quintile and a hypothetical redistribution at the continental scale. A) ITNs distributed in urban areas of sub-Saharan Africa by wealth quintile, 2011-2021, in millions. B) Hypothetical ITN coverage in urban slums of sub-Saharan Africa if all ITNs in urban areas were distributed to urban slums. Included countries are those in Figure S1.

A hypothetical assessment was performed to determine if past ITN distributions in urban areas would have been sufficient to cover people living in urban slums alone (Figure 5B).

Accumulated across urban areas of sub-Saharan Africa, we estimate that between 2015 and 2020, there would have been sufficient ITNs to cover people living in urban slums, assuming one net per two people and that nets could be used for two years^13^. The drop in hypothetical coverage in 2021 could be due to the effects of the COVID-19 pandemic.

Given that reallocating resources away from those with very low risk to provide increased ITN coverage or interventions for those at higher risk would be beneficial for malaria control, we devised two simple hypothetical within-country reallocation schemes of resources spent on ITNs for urban areas over the period 2011-2021 to better target higher risk populations (Figure 6, Table S2).

**Figure 6.**
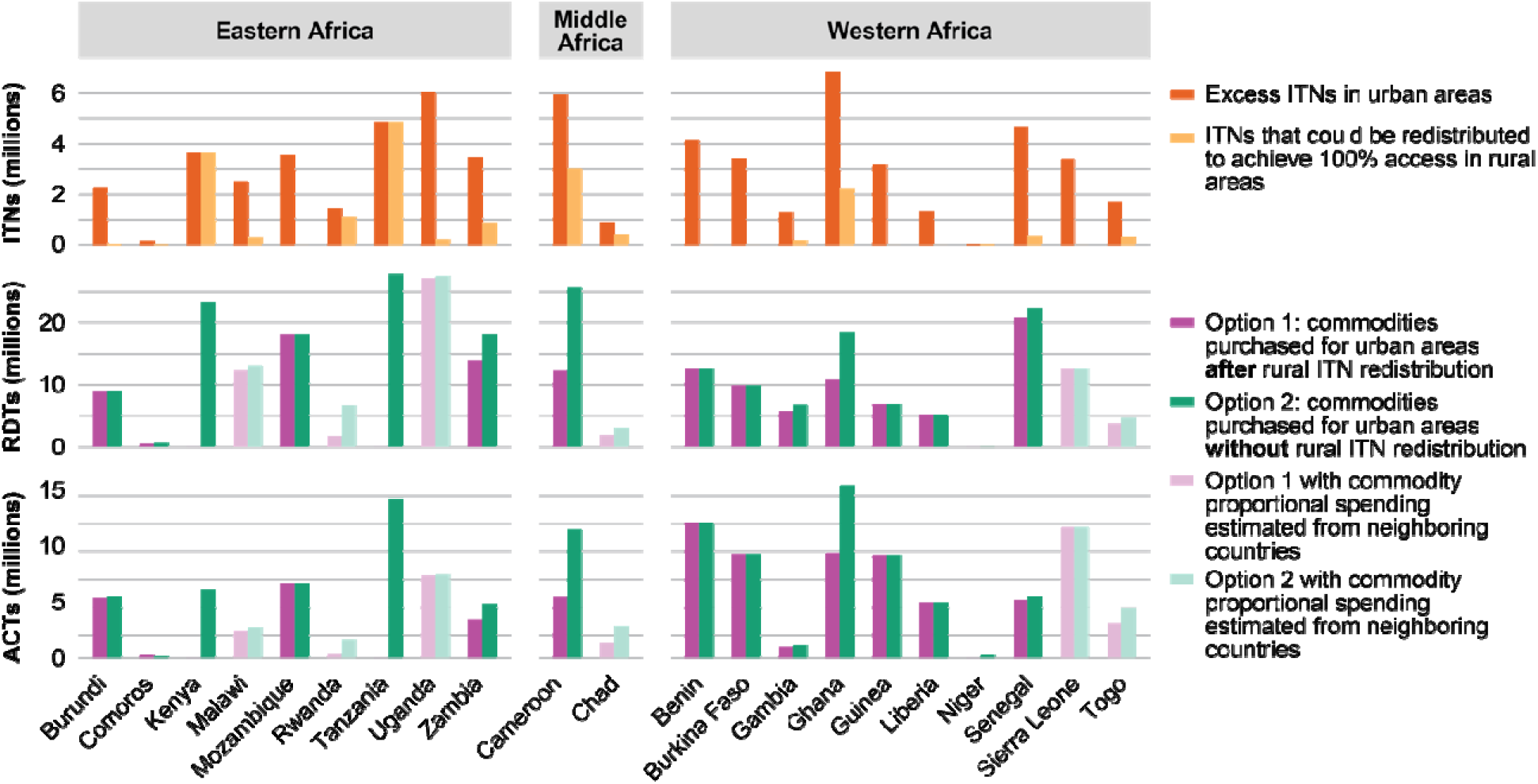
Number of ITNs that could be redistributed (top) and number of RDTs (middle) and ACTs (bottom) that could hypothetically be purchased for urban areas using funds remaining after rural ITN redistribution (Option 1) or all funds from excess nets (Option 2). All values are cumulative over the study period.

In both schemes, ITNs in urban areas were first prioritized to residents of slums (1.0B total ITNs across the 40 countries and 11 years included in this analysis). After allocating ITNs first to slum residents, 21 countries (Figure S8) had at least one year where excess urban ITNs were available for further reallocation (64.9M total ITNs, equivalent to $234M USD). In practice, these funds could be reallocated broadly across many malaria control activities, but we provide two simple reallocation options to illustrate what might be possible with savings of this magnitude.

Under reallocation option 1, the 64.9M remaining excess urban ITNs were redistributed to rural areas within the same country until rural need was met, with rural need defined as 100% access. This resulted in redistribution of 17.5M ITNs to rural areas, with 47.4M ITNs remaining, equivalent to $166M USD. Countries where the most ITNs could have been distributed to rural areas were Kenya, Tanzania, Cameroon, and Ghana. The $166M remaining funds would be able to purchase a total of 185M RDTs and 98M ACT courses, for example for residents of urban slums and informal settlements.

Alternatively, under reallocation option 2, the 64.9M excess urban ITNs were converted entirely to funds, which could have purchased 272M RDTs and 137M ACT courses.

## Discussion

Over $1.0B U.S. dollars were spent on over 440M ITNs in urban areas of sub-Saharan Africa between 2011 and 2021. Malaria prevalence in urban areas is generally low, especially in large cities, and much lower than in rural areas. While malaria prevalence in urban areas was higher for those in the lowest wealth quintiles, ITN access was not necessarily so as well. We found little evidence that ITN access was associated with decreasing risk of malaria infection in urban areas at the continental scale. A theoretical exercise estimated that limiting public ITN distributions in urban areas to slum residents only could have allowed more ITNs to be distributed to areas of higher need or freed up funds for the implementation of other effective interventions wherever they are most needed.

Other studies have also found household wealth and modern housing to be associated with lower risk of malaria^14,15^. In Tanzania, ITNs provided less protection against malaria in Dar es Salaam compared to rural areas, likely due to mosquito-proofing the homes and the higher proportion of outdoor biting in Dar es Salaam^6^. This study found no association between urban malaria prevalence and ITN use, which is complex, depends on many factors, and is subject to respondent bias. People are less likely to sleep under an ITN when they perceive lower risk of malaria^16,17^, as can be the case in urban centers compared with rural areas^18^.

ITNs are most effective in areas with locally-acquired malaria rather than imported cases and where indoor night-time biting is common. Travel history data and entomological surveillance are critical to understanding where these conditions are met. Urban ITN distribution campaigns could target urban sub-populations at highest risk for malaria, such as people living in slums and/or areas near agricultural fields^19,20^. Focusing ITN distribution campaigns in urban slums need not preclude other residents from obtaining nets. Wealthier households could purchase ITNs from retail stores, if desired^21,22^. A multi-country study from 2014-2017 found that 3-40% of urban households in sub-Saharan Africa (varying by country) had purchased at least one net^21^.

Urban areas are not all the same. Malaria prevalence in urban areas outside the country’s largest urban center was often much higher than in the largest center. Even in the regions where large cities are located, urban areas can be immediately abutted by rural areas, and understanding the urban extent in these large cities are crucial. Urban strategies should therefore be tailored to each urban area’s individual epidemiological context rather than assume that all urban areas in a country have similar risk and determinants of transmission.

There are several limitations to this analysis, which aims to paint an approximate overall picture of the allocation of ITNs in urban areas across a continent rather than understand deeply the local epidemiological and intervention context. DHS surveys are conducted only every few years, leaving large gaps between survey years in some countries. Within urban areas, DHS is undersampled^23-25^, which means analysis results at that level are uncertain, although the consistency of patterns across countries and surveys is reassuring. It was not possible to distinguish for all countries whether ITNs used were acquired through programmatic distribution or purchased by individuals. ITN procurement costs were estimated from publicly available documents from donors. The reported costs are estimates based on previous years and expected need and may not be the actual amount of funds used to purchase ITNs. Twenty-five countries did not have any ITN cost estimates publicly available, so the ITN procurement cost was approximated from neighboring countries. ITN distribution costs were not found in donor documents for half of the countries of study, and therefore, distribution costs were not included in this analysis. Additionally, ITN use was determined from the DHS by asking the household members if they slept under a net the night before the survey; therefore, the consistency of ITN use was unknown.

Prior to early 2025, there was already an annual gap of $3.7B USD in funding for malaria control and elimination efforts^12^. Uncertainty in the future of global aid for malaria, including the $800 million annual investment of the President’s Malaria Initiative and other US commitments to global health, mean that the need to prioritize is more urgent than ever. Malaria programs should carefully consider whether reprioritization of resources away from ITNs in less vulnerable parts of urban areas, or urban areas entirely, may be a necessary strategy. It is imperative that national malaria programs have a sufficient understanding of malaria transmission patterns and populations most at risk within their countries, especially in the context of insufficient funding for malaria. Programs need to strengthen case-based surveillance in urban areas to enable reprioritization at a finer scale. Decision-making regarding mass ITN distribution campaigns in urban areas should be informed by local data on risk, vulnerability, travel, and vector behavior to the extent possible to provide ITNs to those at highest risk of malaria. WHO already has a framework for prioritization in urban areas^2^, which can be further refined as countries test these ideas in practice.

## Methods

### Estimating the number of nets distributed in urban and rural areas

A total of 87 surveys covering 31 countries were included. To estimate the number of insecticide-treated nets (ITNs) distributed in urban and rural areas of sub-Saharan Africa between 2011 and 2021, we calculated the proportion of ITNs found in urban and rural areas from Demographic and Health Surveys (DHS) and Malaria Indicator Surveys (MIS) (here referred to collectively as DHS) over the same time frame, accounting for survey weights, then multiplied by the total number of ITNs distributed. The population of each country by urban vs. rural for each year of analysis was estimated from total population estimates from the United Nations and the proportion of the population living in urban areas estimated at 5-year intervals from the World Urbanization Prospects 2018 Source^26-27^. A linear interpolation of the proportion of people living in urban areas by year was estimated from the 5-year intervals between 2010 and 2020 from the UN source dataset.

DHS employs a two-stage stratified sample design to provide representative estimates at the national and urban/rural administrative levels^28,29^. The number of ITNs owned by households was extracted separately for urban and rural households according to country-specific definitions of urban and rural used in the DHS. Survey weights were applied via the R Survey package^30^ to calculate, for each year of DHS available, the proportion of a country’s ITNs that were in urban areas. A linear interpolation was used to estimate the proportion of nets in urban areas for years between surveys. The proportion of nets in urban and rural areas were each multiplied by the total number of ITNs distributed by year for each country from the 2022 World Malaria Report^31^ and WHO repositories (personal communication).

To estimate the number of ITNs in urban areas by wealth quintile, the proportion of ITNs by wealth quintile was estimated from the DHS.

### Net procurement costs

Total yearly ITN procurement cost was extracted from the countries’ President’s Malaria Initiative (PMI) Malaria Operational Plans (MOPs) and the Global Fund’s Malaria funding reports^32^ (Figure S9). Twenty-five countries had cost data directly estimated from the MOPs and Global Fund reports, and 18 countries’ unit cost per ITN was estimated from their neighboring countries. Procurement unit cost per ITN was estimated yearly for 43 countries for the period 2011-2021 (Figure S10). Some countries reported procurement costs for standard ITNs and nets treated with piperonyl-butoxide (PBO), whereas others reported only one ITN type. Therefore, different analytic methods were used to analyze the data. If both ITN and PBO costing data was available, a weighted average of the number of ITNs and PBOs was used to calculate the ITN unit cost per year. If a country had no costing data available (n= 18), the average unit cost per ITN from the neighboring countries was used. All cost data were adjusted for inflation to 2021 US dollars value. Total cost spent on nets (Figure 1E) was calculated by multiplying the number of nets distributed in a year times the unit cost for that year. Three of the 43 countries were dropped from the total cost estimation since they did not have an estimated number of nets distributed available.

### Urban malaria prevalence, ITN access, ITN use, and associated factors

Sixty-four DHS and MIS conducted between 2010-2021 were included from 23 countries (Figure S1, Table S1). Countries were included in this analysis if they tested a subset of children 6-59 months—heretofore referred to as ‘children under 5’, for malaria using rapid diagnostic tests (RDT). Prevalence by RDT was used since all countries tested for malaria by RDT but not necessarily microscopy. The age of interest (children under 5) is motivated by data availability of statistically representative georeferenced data. The average malaria prevalence among children 6-59 months in urban areas, within the largest urban center, and in all urban areas excluding the largest urban center was extracted. The largest urban center was defined as the most populous area that is considered an urban center by the Global Human Settlement Layer^33^. Household wealth was a binary variable defined as wealthy for the highest and second highest wealth quintiles, and not wealthy for the lower three wealth quintiles.

ITN access was defined as the number of de facto household members who could sleep under an ITN, assuming the ITN was used by up to two people, divided by the number of persons who slept in the household the night before the survey^34^. ITN use was defined as the proportion of persons who slept under an ITN the night before the survey. The average malaria prevalence, ITN access, and ITN use was calculated for each binary wealth category. ITN access and use for household members was calculated for urban clusters, accounting for the survey design.

Cluster-level ITN access, ITN use, proportion with modern housing, proportion living in a wealthy household, precipitation, enhanced vegetation index (EVI), and relative humidity were assessed as potential factors affecting malaria prevalence in urban areas. Data were analyzed at the DHS cluster-level and only clusters located in urban areas were included (7,478 clusters). A household was deemed to have modern housing if it had an improved floor, roof, and walls (Table S3). Household wealth was a binary variable defined as wealthy for the highest and second highest wealth quintiles, and not wealthy for the lower three wealth quintiles.

Cluster geospatial coordinates were obtained from DHS. Since DHS displaces the location of urban clusters by up to 2 km for confidentiality purposes, raster values were aggregated across a 2km buffer. Precipitation, EVI, and relative humidity were extracted for each DHS cluster from raster maps obtained from publicly available sources^35-37^ (Table S4). Environmental variables were averaged by month (or summed for the entire month for total precipitation) and were lagged by two months^38,39^. If the cluster had more than one survey month, the environmental data was lagged from the first survey month in the cluster^40^. For the six surveys (Mali 2012, 2015, 2018, 2021; Rwanda 2017; and Gambia 2013) without cluster-level GPS coordinates available from DHS, environmental data were extracted from the lowest-level administrative unit of the cluster.

Descriptive statistics were calculated taking into account the survey design using the R survey package^30^.

### Multivariable modeling

Exploratory analyses and review of the literature informed variable selection for the multivariate modeling. Correlations using Kendall’s Tau statistic were assessed between each covariate of interest and the outcome (test positivity rate).

A zero-inflated Poisson regression was used to model malaria test positivity rate controlling for the factors described above and survey year. Year was included as a continuous variable. The zero-inflated model was selected because the majority of clusters (61%) had no children positive for malaria. The dependent variable was the number of children 6-59 months who tested positive for malaria, adjusted for the number of children tested per cluster using the offset term. The glmmTMB package was used to conduct the modeling. The models were fit using a random intercept for country to account for variation between countries. Model diagnostic results including the Q-Q plot and the residuals vs. predicted values plot are included in Figure S7. All analyses were conducted in R version 4.2.3 (R Foundation for Statistical Computing, Vienna, Austria).

### Urban ITN redistribution scenarios

We considered a hypothetical strategy where ITNs distributed in urban areas were reserved for persons living in urban slums and any remaining nets were considered excess nets. Countries with sufficient data to calculate ITN funds and the proportion of funds used for RDTs and ACTs were included (n=15, Figure S8). Hypothetical reallocations were performed for each country and year.

Urban ITN need was calculated from the population living in urban slums divided by two, assuming two persons can sleep under one ITN^29^. The proportion of urban populations living in slums and the total urban population was obtained from the UN Habitat Urban Indicators Database^41^, which defines slum households as those with lack of access to improved water, lack of access to improved sanitation, lack of sufficient living area and quality/durability of structure. Excess ITNs were then calculated by subtracting the total number of nets distributed in urban areas by this approximation of urban need. The number of excess urban ITNs per country was multiplied by the unit ITN cost to provide an estimate of the available funds if the excess ITNs had not been purchased. A flowchart of this process is provided in Figure S11. Following these steps, 21 countries had at least one year between 2011-2021 where there were hypothetically enough ITNs in urban areas to provide sufficient ITNs for those living in slums with some excess ITNs remaining.

For each year with excess urban ITNs, we assessed two options for repurposing the excess ITNs. In Option 1, excess ITNs from urban areas were first redistributed to rural areas to fulfill 100% of rural need (defined as the rural population divided by 2), using rural access from DHS as the baseline. Any further excess ITNs were converted back to funds using the ITN unit cost, and these funds were allocated to purchase commodities (RDTs and ACTs) for urban areas. In Option 2, all excess ITNs from urban areas were converted to funds using the ITN unit cost, and these funds were allocated to purchase RDTs and ACTs for urban areas.

The proportion of the excess funds that would go towards purchasing RDTs vs. ACTs was determined on a country-specific basis, as the ratio of RDTs to ACTs needed can depend on local factors such as transmission intensity. For each country, the number of ACT treatment courses delivered and RDTs distributed in 2021 (obtained from World Malaria Report 2022, Annex 4D) were multiplied by their respective unit costs (estimated at the yearly level) to calculate the proportional allocation of funds used for RDTs and ACTs. The annual unit costs of ACTs and RDTs (2011-2021) were estimated from PMI’s MOPs for each country^32^ and were linearly interpolated for years where there was no data available. The funds for RDTs were calculated by multiplying the estimated unit cost of RDTs (in 2021) by the number of RDTs distributed in 2021, and the funds for ACTs was calculated in the same manner. These two commodity costs were summed for each country and the respective proportions of funds devoted to each of RDTs and ACTs were calculated. This proportional allocation was then used for all years of hypothetical reallocation. Estimated number of commodities (RDTs and ACTs) that could be purchased was calculated by multiplying the respective yearly funds available for redistribution by the proportional allocation of funds for RDTs and ACTs.

## Data Availability

All data produced in the present work are contained in the manuscript

## Acknowledgements

CL, AS, OOD, and JG were supported by a grant from the Gates Foundation to JG (INV-048410). IDO, CC, and NSA were supported by a grant from the Gates Foundation to IDO (INV-036449). AN was supported by a grant from the Gates Foundation to AN (INV-061337).

## Disclaimer

Beatriz Galatas is a staff member of the World Health Organization. This author alone is responsible for the views expressed in this article and does not necessarily represent the decisions, policy or views of the World Health Organization.

## Author contributions

AN, IDO, BG, and JG conceptualized the study. CL, AS, CC, and NSA assembled the data. CL, IDO, AS, CC, NSA, and OOD analyzed the data. All authors interpreted the analysis. CL and JG wrote the first draft of the manuscript. All authors reviewed, edited, and approved the final manuscript.

## Availability of code and materials

Analysis code for this work can be found at https://github.com/numalariamodeling/urban-malaria-Africa-publication-2023. All datasets used in this work were publicly sourced as described in the Methods section, supplemented by a few countries’ net distribution numbers shared directly by WHO. Interested parties may contact the Global Malaria Programme at WHO for those data.

