## Supplemental Information for "Efficiencies in the allocation of insecticide-treated nets for malaria prevention in urban Sub-Saharan Africa"

**Figure S1.** Countries included in the urban malaria prevalence, ITN access, ITN use, and associated factors analyses.

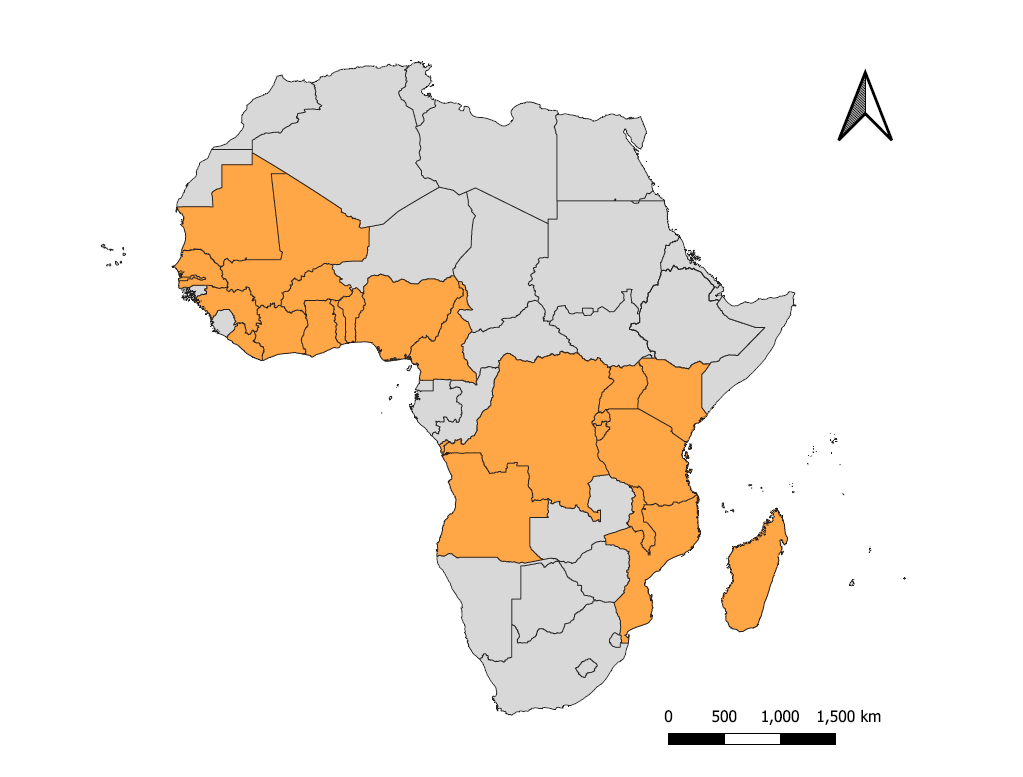

**Figure S2.** Total insecticide-treated nets distributed and proportion in urban vs. rural areas per country.

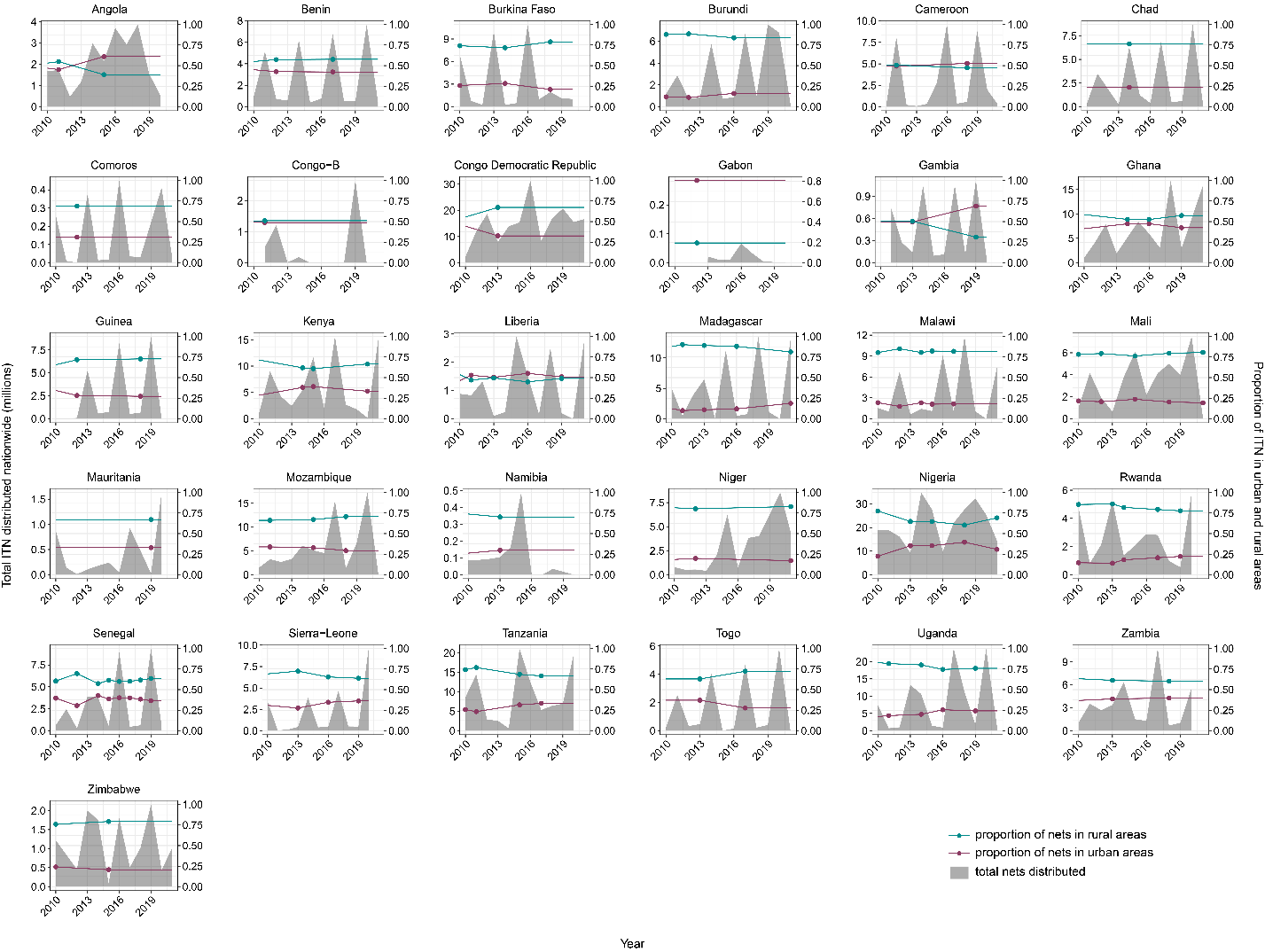

**Figure S3**. Proportion of ITNs in urban areas by the proportion of the country’s population living in urban areas. The red dashed line shows the 1:1 ratio.

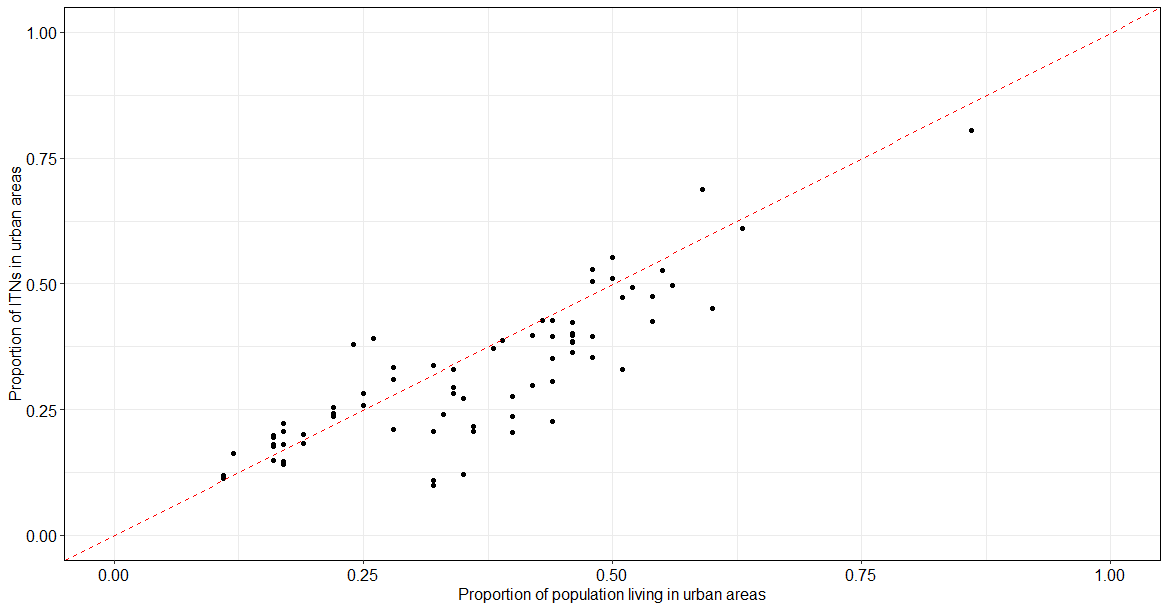

**Figure S4. ITN access and use in urban areas by country.** Shown: all urban areas combined and all wealth strata.

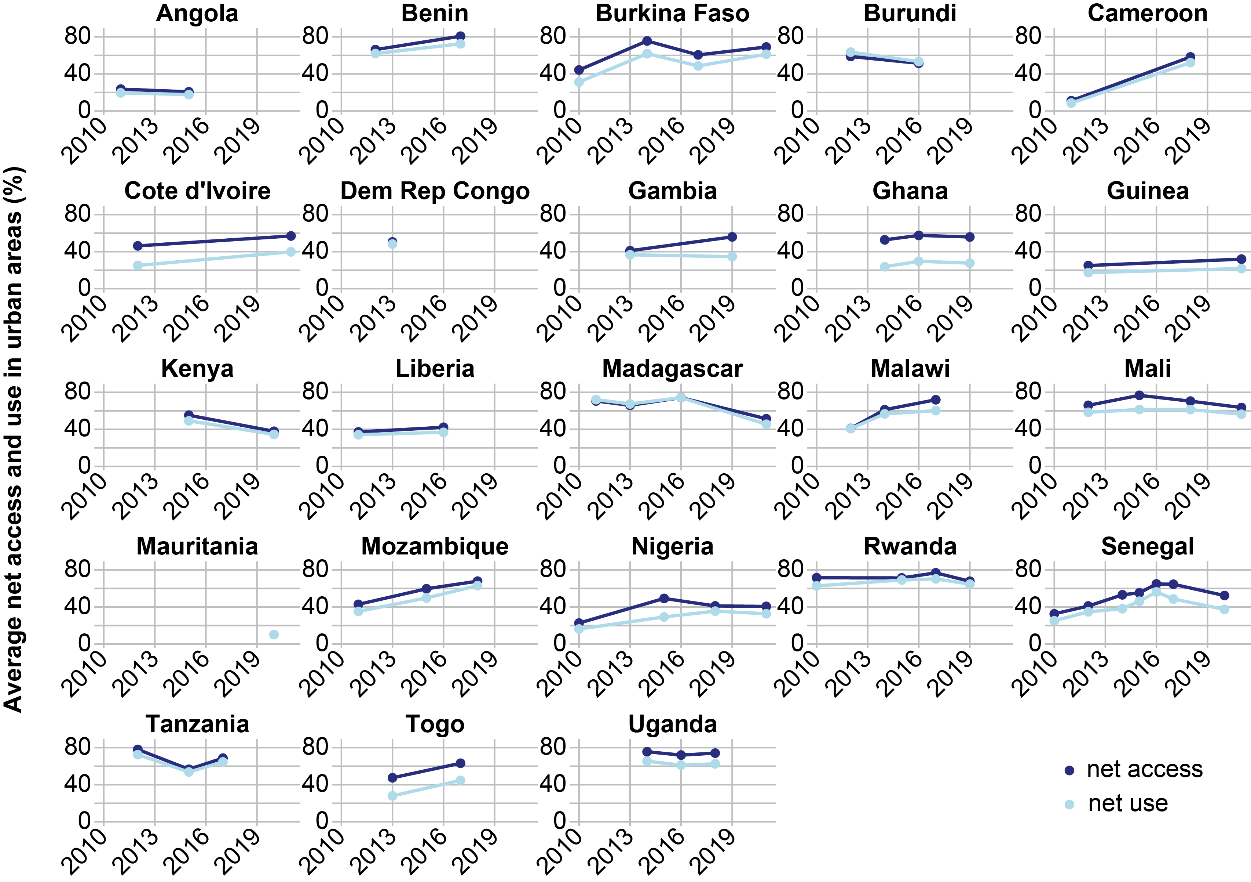

**Figure S5.** Average net access and use in the largest urban center (A), urban areas excluding the largest urban center (B), and rural areas (C).

**A.**

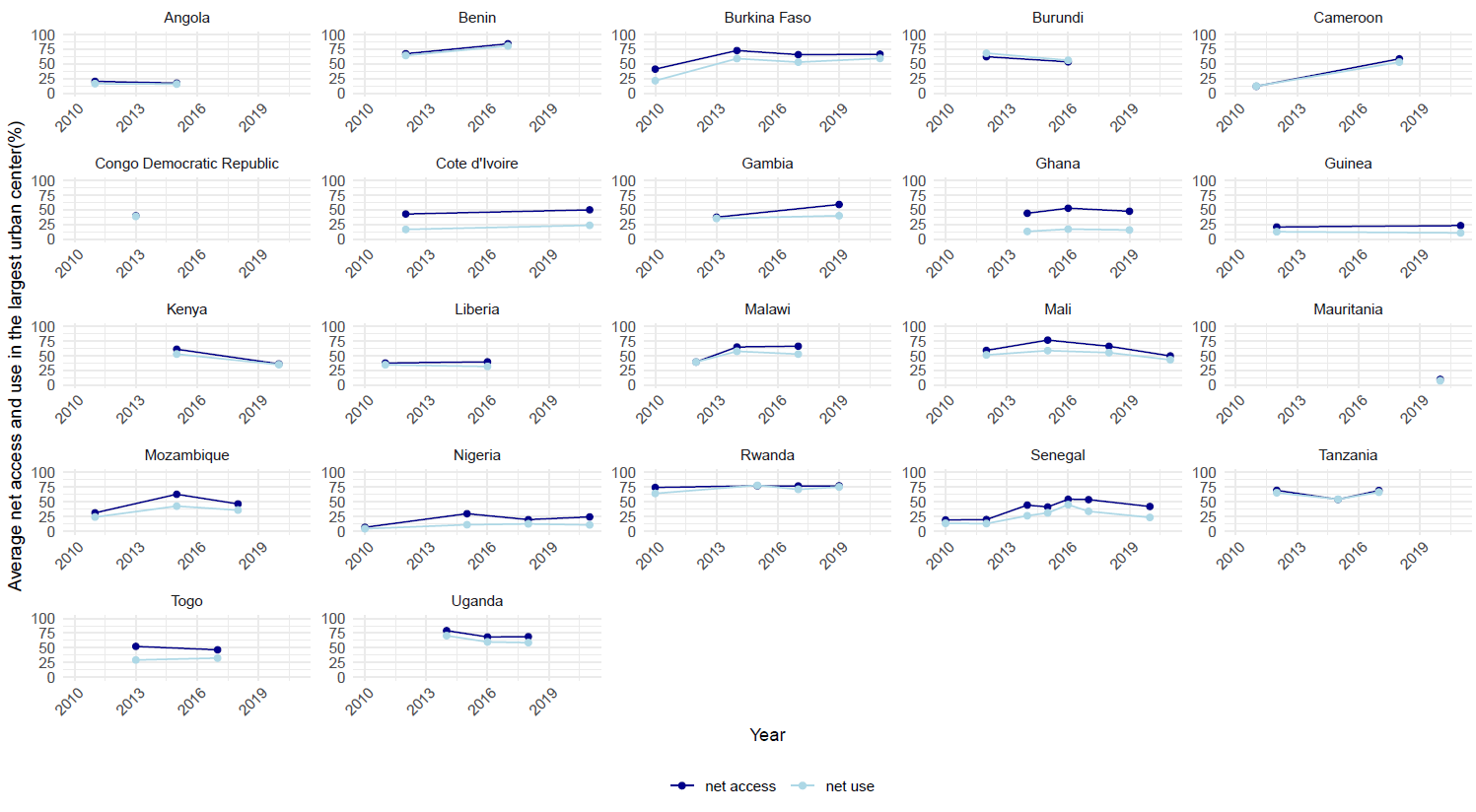

**B.**

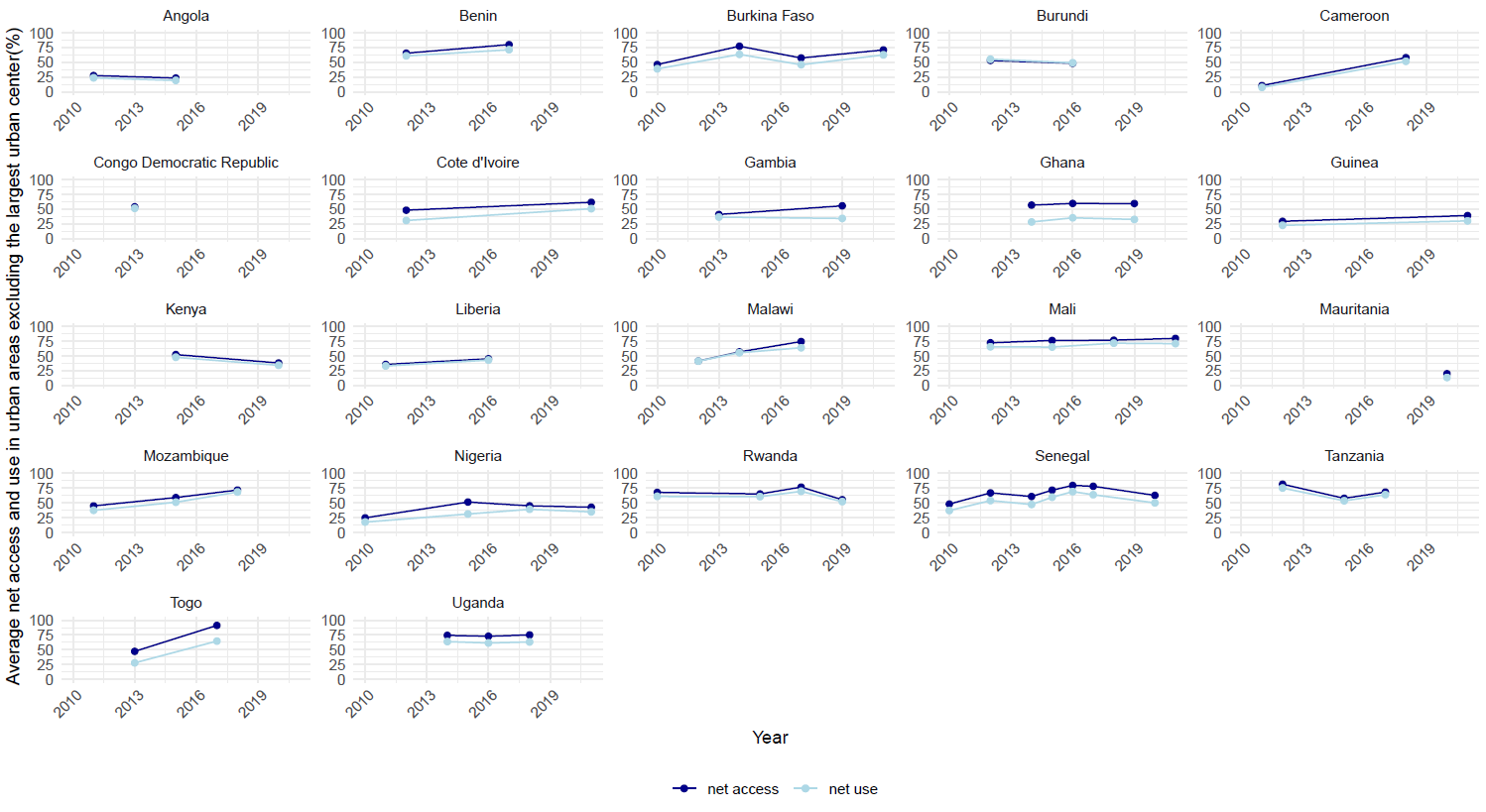

**C.**

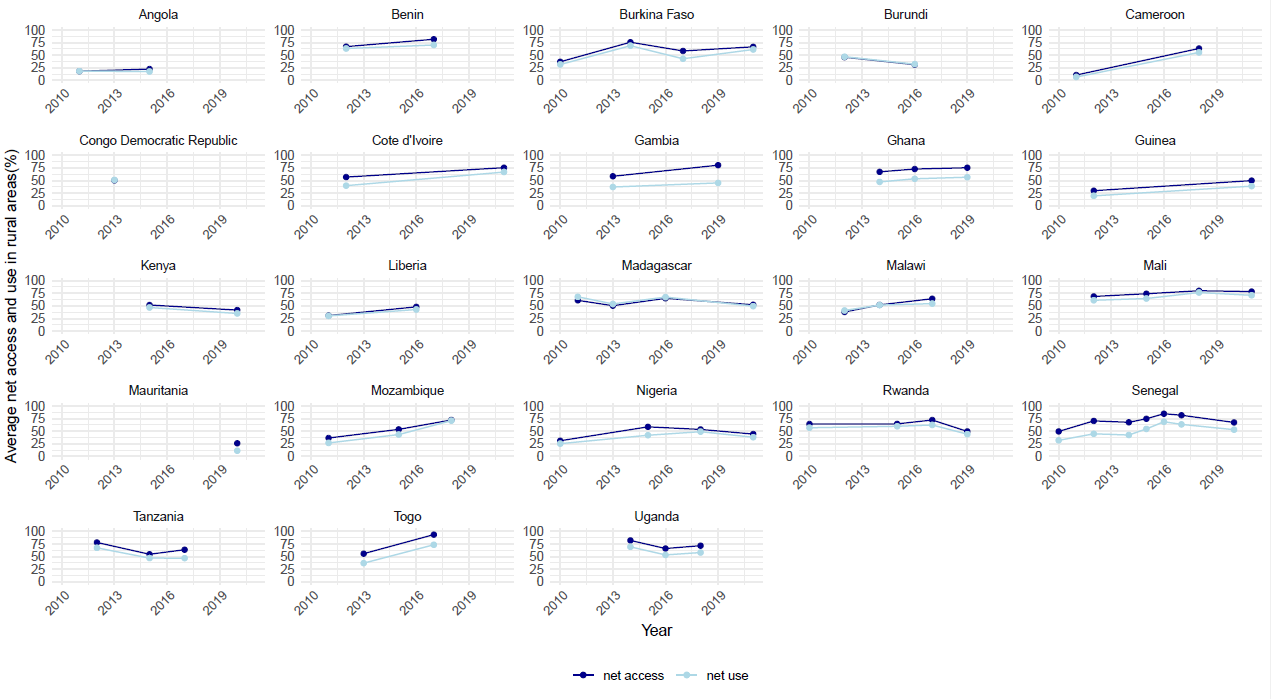

**Figure S6.** ITN use in urban areas, defined as the proportion of the population that slept under an ITN the night before, stratified by wealth quintile.

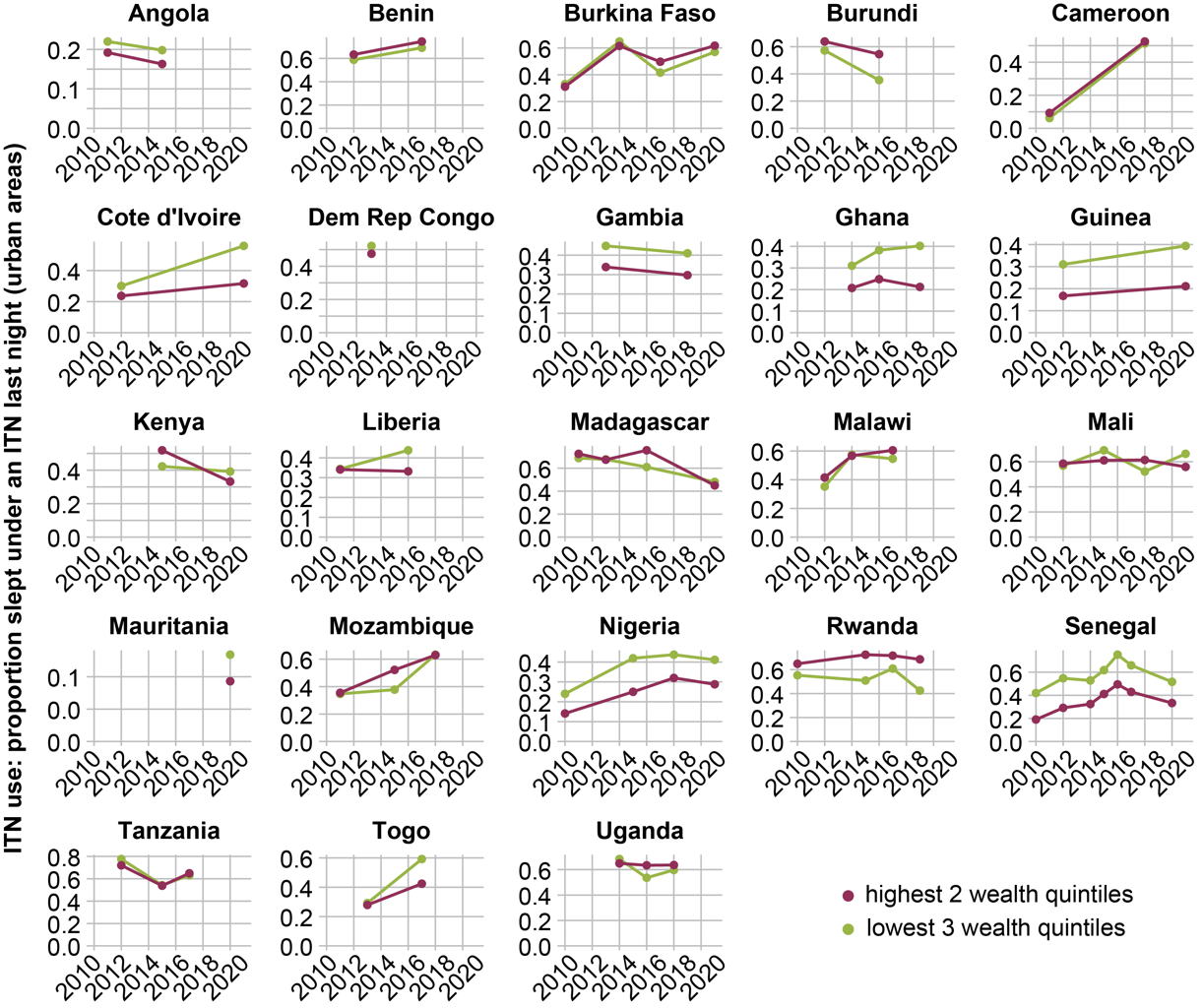

**Figure S7.** A) Diagnostic results from the multivariate modeling. B) Expected urban malaria test positivity rate from zero-inflated Poisson model from 2010-2021. The blue line is the model predicted rate and the shading represents the upper and lower confidence limits (95% CI).

**A**

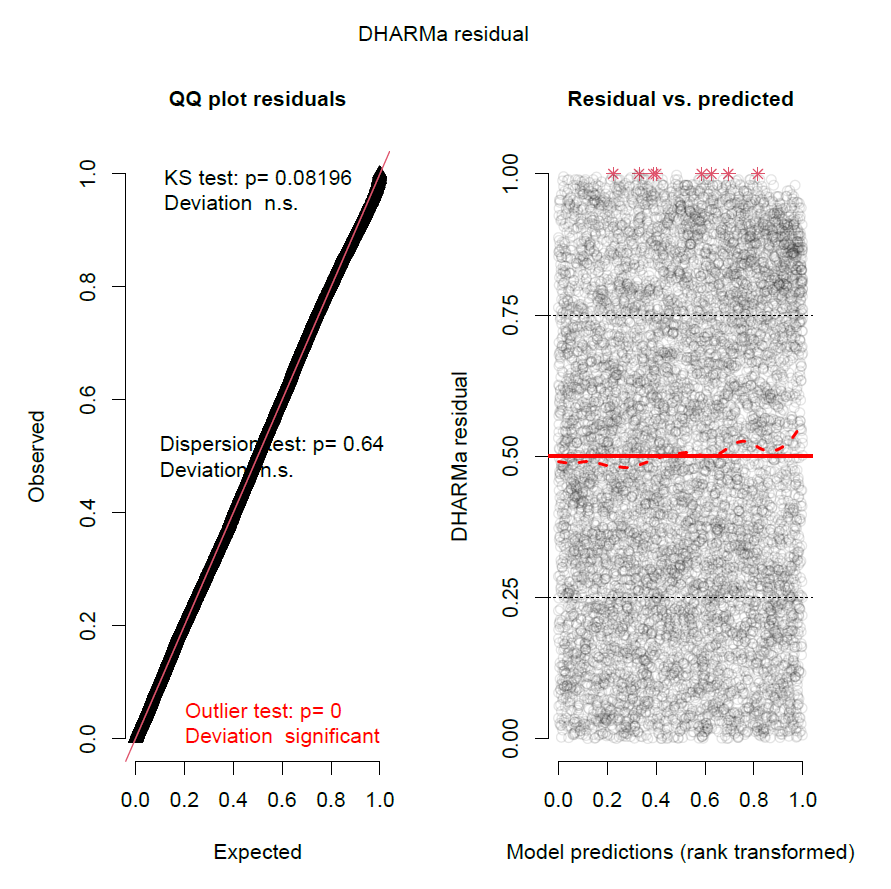

**B**

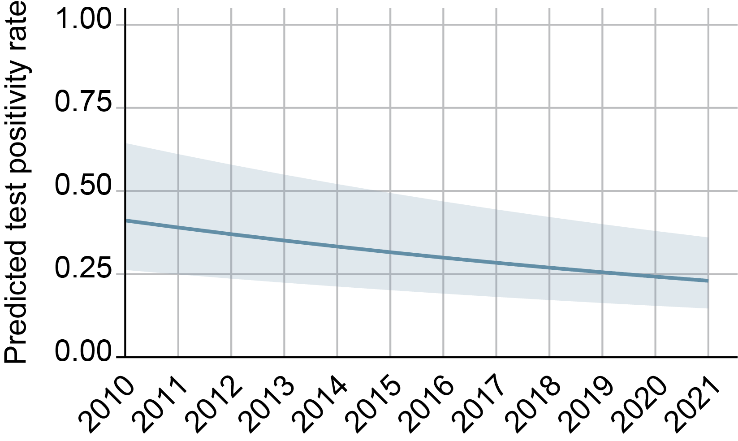

**Figure S8.** Countries with at least one year of excess nets in the insecticide-treated nets funds redistribution analysis. Countries in dark gray did not have an estimation for proportion of funds spent on RDTs vs. ACTs (n= 6).

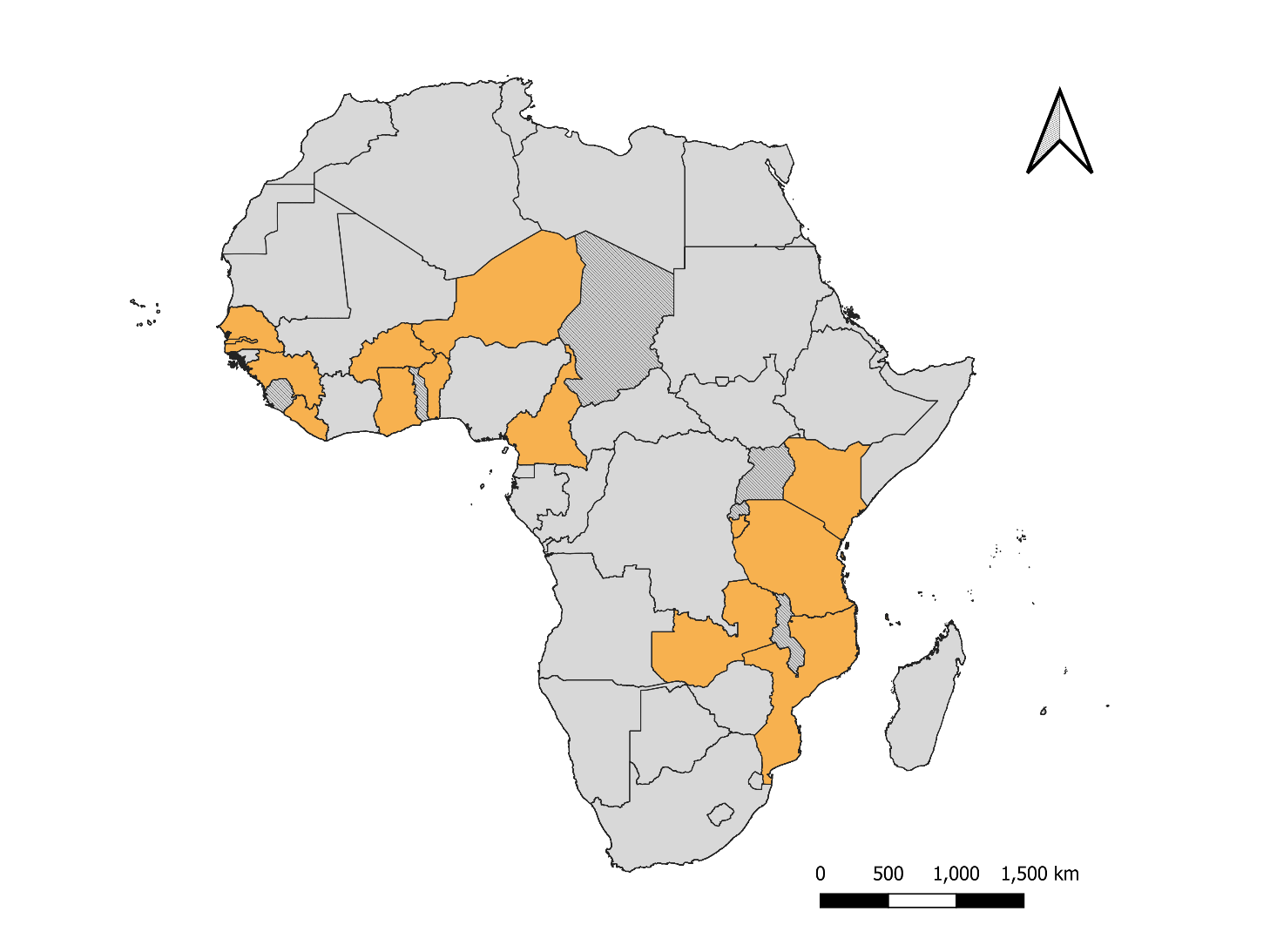

**Figure S9.** Approach for estimating yearly average procurement unit cost of nets per country.

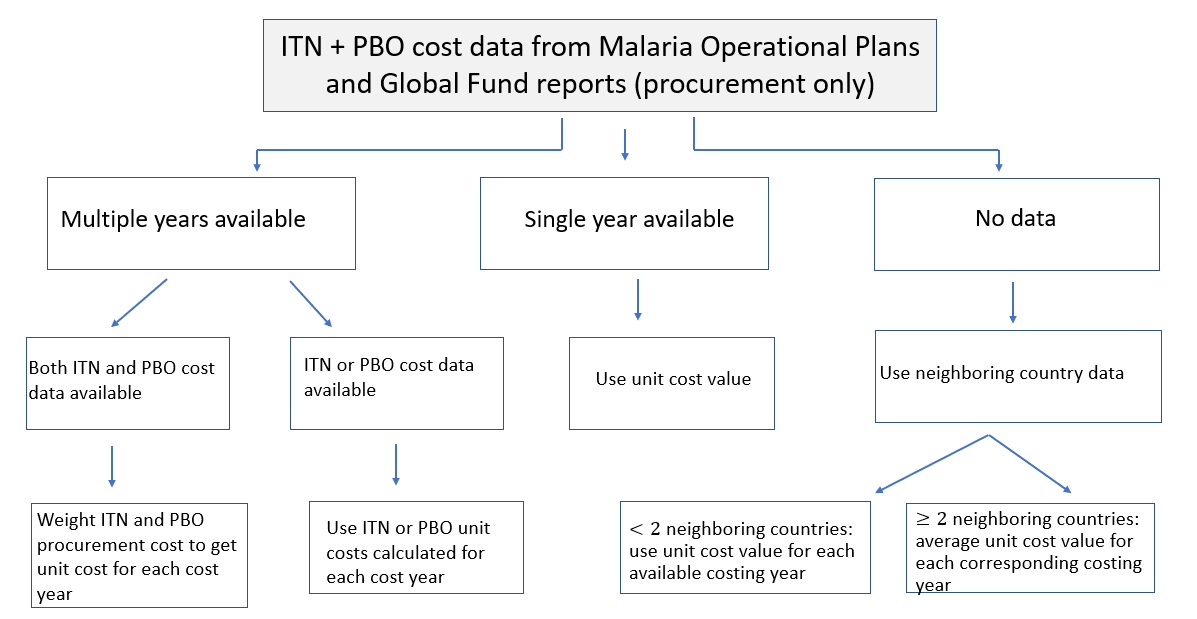

ITN= insecticide-treated net

PBO= piperonyl-butoxide treated net

**Figure S10.** Procurement unit cost of insecticide-treated nets (ITNs) and pyrethroid-piperonyl butoxide (PBO) nets in sub-Saharan Africa from continuous distribution and mass distribution campaigns from 2012- 2021.

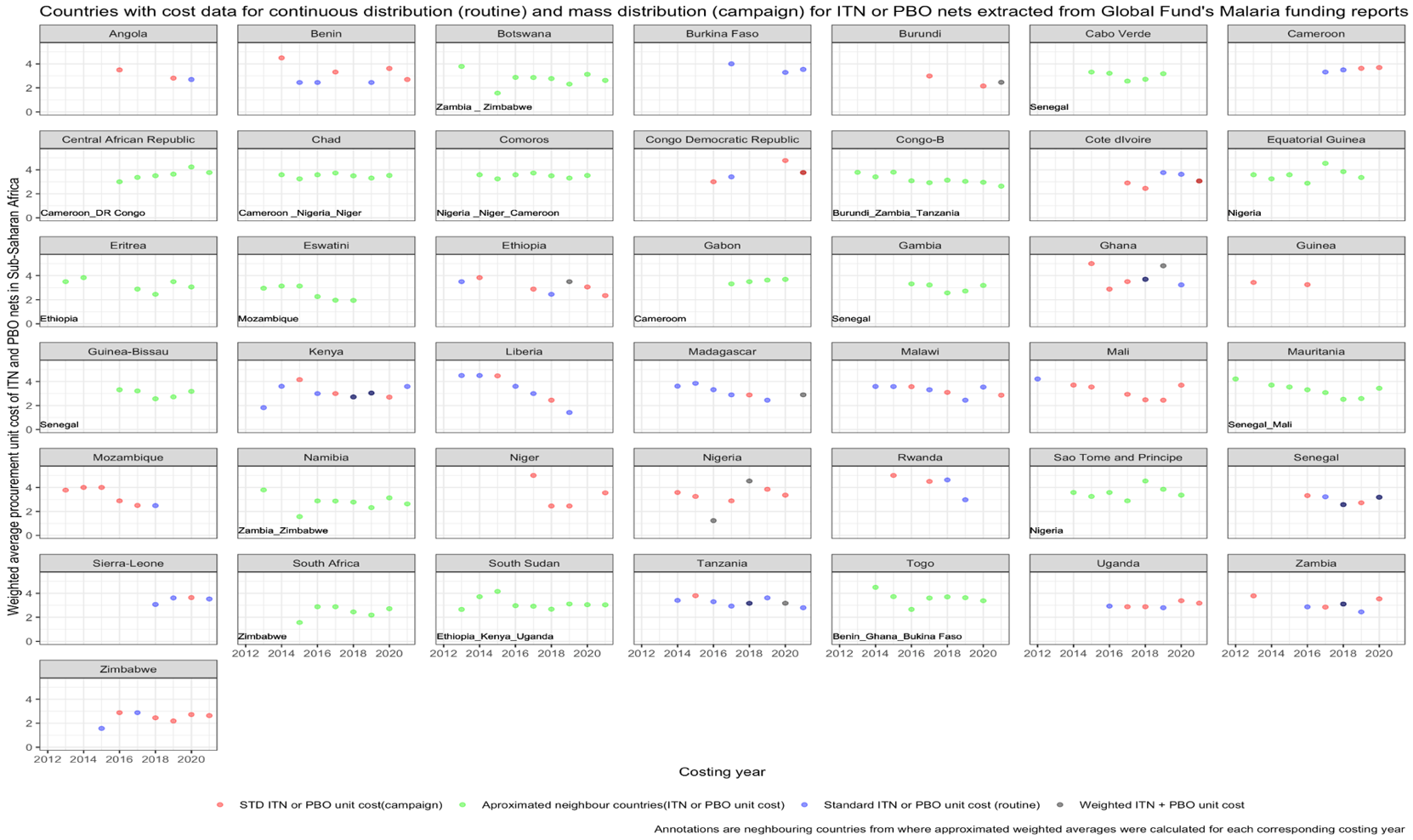

**Figure S11.** Flowchart of calculation of number of excess urban insecticide-treated nets in urban areas that could be rerouted to provide nets to rural areas, or purchase RDTs and ACTs for urban areas.

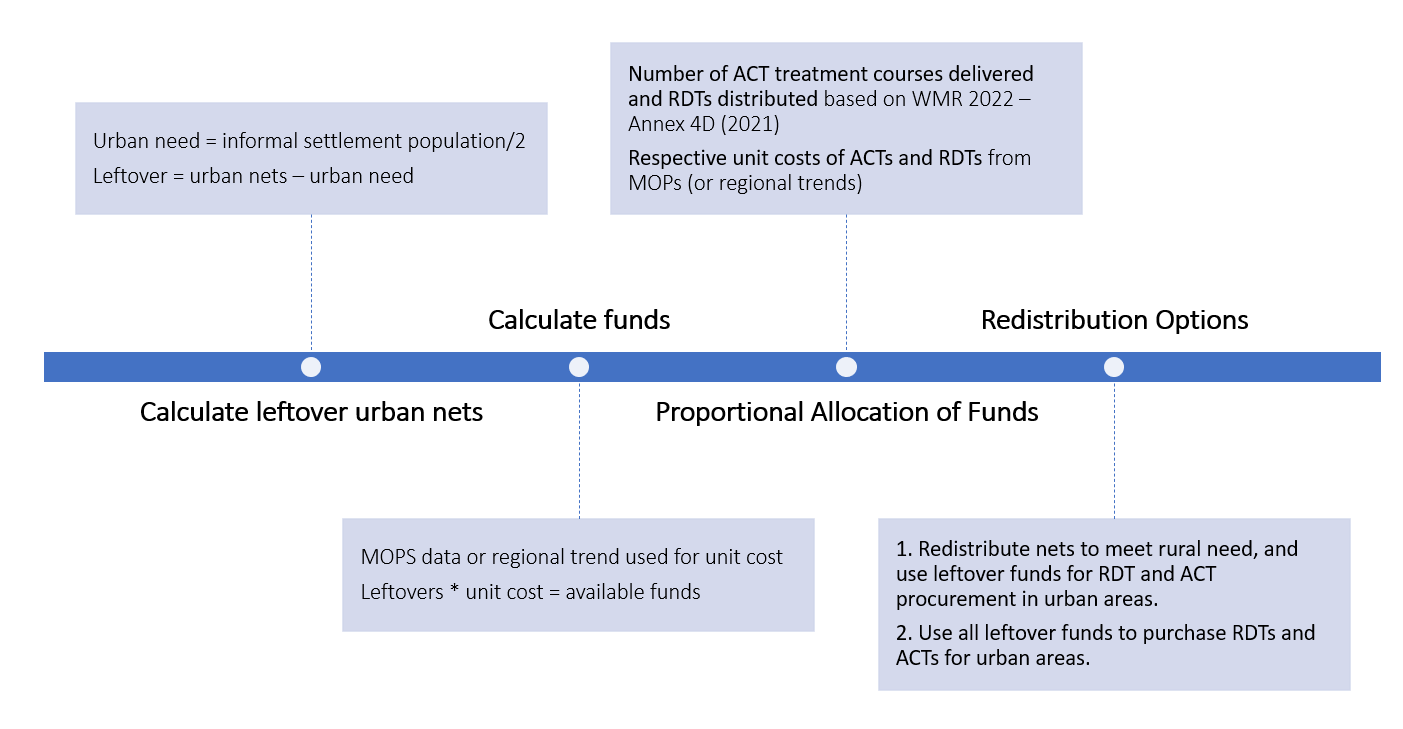

Rural need= rural population/ 2

**Table S1**. Number of countries included in each analysis.

| **Analysis** | **Inclusion criteria** | **Number of countries included** |
| --- | --- | --- |
| Number of nets in urban and rural areas | - At least one DHS/MIS with the proportion of ITNs in urban and rural areas between 2011- 2021 - Annual number of ITNs distributed per country obtained from national malaria programs | 31: Angola, Benin, Burkina Faso, Burundi, Cameroon, Chad, Congo, Democratic Republic of the Congo, Eswatini, Gabon, Gambia, Ghana, Guinea, Kenya, Liberia, Madagascar, Malawi, Mali, Mauritania, Mozambique, Namibia, Niger, Nigeria, Rwanda, Senegal, Sierra Leone, Tanzania, Togo, Uganda, Zambia, Zimbabwe |
| Net procurement costs | - Annual number of ITNs distributed per country obtained from national malaria programs - ITN costing data available from MOPs | 40: **Angola**, **Benin,** Botswana, **Burkina Faso, Burundi, Cameroon**, Cape Verde, Central African Republic, Chad, Comoros, Congo, **Democratic Republic of the Congo**, Eritrea**, Ethiopia**, Eswatini, Equatorial Guinea, Gabon, Gambia**, Ghana, Guinea,** Guinea-Bissau**, Kenya, Liberia, Madagascar, Malawi,** **Mali,** Mauritania, **Mozambique,** Namibia, **Niger**, **Nigeria, Rwanda**, **Senegal,** **Sierra Leone,** South Sudan, **Tanzania**, Togo, **Uganda,** **Zambia, Zimbabwe**  24 (with their own cost estimates, not approx. from neighboring countries) (bolded above) |
| Malaria prevalence and associated factors, net access and net usage | - Had at least one DHS/MIS survey between 2010- 2021 - Tested children under five for malaria by RDT | 23: Angola, Benin, Burkina Faso, Burundi, Cameroon, Cote d’Ivoire, Democratic Republic of the Congo, Gambia, Ghana, Guinea, Kenya, Liberia, Madagascar, Malawi, Mali, Mauritania, Mozambique, Nigeria, Rwanda, Senegal, Tanzania, Togo, Uganda |
| Urban ITN redistribution options | - Annual number of ITNs distributed per country and ITN unit cost data available - Had at least one year with excess ITNs in urban areas in a mass distribution year | 21: **Benin**, **Burkina Faso**, **Burundi**, **Cameroon**, Chad, **Comoros**, **Gambia**, **Ghana**, **Guinea**, **Kenya**, **Liberia**, Malawi, **Mozambique**, **Niger**, Rwanda, **Senegal**, Sierra Leone, **Tanzania**, Togo, Uganda, **Zambia** |
|  | - Number of ACT and RDTs distributed in 2021 and unit cost data available (for estimating the proportion of funds used for RDTs v. ACTs) to estimate number of RDTs and ACTs that could be purchased | 15 (bolded above) |

**Table S2.** Redistribution summary: number of excess urban nets, and number of RDTs and ACTs that could be procured for urban areas using funds from excess nets in urban areas (Option 1), and funds from excess nets after redistribution to meet rural net need (Option 2). Only country-year pairs were there were excess nets in urban areas.

| **Country** | **Year** | **Urban slum population (M)** | **Excess nets from urban areas** | **Net funds available for urban areas (Option 1)** | **# RDTs for purchase in urban areas (Option 1)** | **# ACTs for purchase in urban areas (Option 1)** | **Nets needed in rural areas** | **Excess nets remaining** | **Net funds remaining for urban areas (Option 2)** | **# RDTs for purchase in urban areas (Option 2)** | **# ACTs for purchase in urban areas (Option 2)** |
| --- | --- | --- | --- | --- | --- | --- | --- | --- | --- | --- | --- |
| Benin | 2011 | 2.88 | 815,561 | 4,013,023 | 2,081,045 | 2,242,291 | - | 815,561 | 4,013,023 | 2,081,045 | 2,242,291 |
| Benin | 2014 | 3.21 | 1,035,091 | 5,093,235 | 2,952,925 | 2,266,571 | - | 1,035,091 | 5,093,235 | 2,952,925 | 2,266,571 |
| Benin | 2017 | 3.58 | 1,074,718 | 3,767,737 | 2,989,843 | 3,210,559 | - | 1,074,718 | 3,767,737 | 2,989,843 | 3,210,559 |
| Benin | 2020 | 3.99 | 1,244,900 | 4,506,323 | 4,489,227 | 4,375,651 | - | 1,244,900 | 4,506,323 | 4,489,227 | 4,375,651 |
| Burkina Faso | 2013 | 2.07 | 1,722,096 | 7,273,866 | 4,948,388 | 4,651,219 | - | 1,722,096 | 7,273,866 | 4,948,388 | 4,651,219 |
| Burkina Faso | 2016 | 1.98 | 1,706,010 | 7,205,921 | 4,902,166 | 4,607,772 | - | 1,706,010 | 7,205,921 | 4,902,166 | 4,607,772 |
| Burundi | 2011 | 0.57 | 50,859 | 160,519 | 228,450 | 138,277 | 50,859 | - | - | - | - |
| Burundi | 2014 | 0.58 | 501,675 | 1,583,362 | 2,253,439 | 1,363,971 | - | 501,675 | 1,583,362 | 2,253,439 | 1,363,971 |
| Burundi | 2017 | 0.59 | 797,141 | 2,515,897 | 3,580,623 | 2,167,295 | - | 797,141 | 2,515,897 | 3,580,623 | 2,167,295 |
| Burundi | 2019 | 0.59 | 930,504 | 2,316,273 | 2,996,836 | 1,797,596 | - | 930,504 | 2,316,273 | 2,996,836 | 1,797,596 |
| Cameroon | 2011 | 5.17 | 1,420,807 | 4,981,066 | 5,402,959 | 2,306,670 | 889,034 | 531,773 | 1,864,290 | 2,022,194 | 863,329 |
| Cameroon | 2016 | 5.18 | 2,377,063 | 8,333,509 | 9,039,352 | 3,859,145 | 738,068 | 1,638,995 | 5,745,990 | 6,232,672 | 2,660,897 |
| Cameroon | 2019 | 5.06 | 2,145,939 | 7,887,883 | 11,175,344 | 5,310,299 | 1,380,978 | 764,961 | 2,811,787 | 3,983,665 | 1,892,957 |
| Chad* | 2014 | 2.56 | 217,407 | 835,079 | 445,380 | 812,561 | 217,407 | - | - | - | - |
| Chad* | 2017 | 2.84 | 211,063 | 743,746 | 720,885 | 770,211 | 211,063 | - | - | - | - |
| Chad* | 2020 | 3.17 | 472,598 | 1,758,882 | 1,880,606 | 1,284,281 | - | 472,598 | 1,758,882 | 1,880,606 | 1,284,281 |
| Comoros | 2013 | 0.14 | 45,989 | 164,761 | 137,131 | 32,341 | 6,114 | 39,875 | 142,857 | 118,900 | 28,041 |
| Comoros | 2016 | 0.16 | 62,180 | 217,006 | 315,164 | 47,568 | - | 62,180 | 217,006 | 315,164 | 47,568 |
| Comoros | 2020 | 0.18 | 39,783 | 117,401 | 161,264 | 40,660 | 23,087 | 16,696 | 49,271 | 67,679 | 17,064 |
| Gambia | 2011 | 0.49 | 119,170 | 541,548 | 620,017 | 185,086 | 33,063 | 86,107 | 391,298 | 447,997 | 133,735 |
| Gambia | 2014 | 0.52 | 290,210 | 1,254,400 | 1,292,549 | 225,411 | - | 290,210 | 1,254,400 | 1,292,549 | 225,411 |
| Gambia | 2017 | 0.56 | 377,265 | 1,361,578 | 2,023,858 | 312,004 | 41,079 | 336,186 | 1,213,320 | 1,803,487 | 278,031 |
| Gambia | 2019 | 0.58 | 478,730 | 1,492,695 | 2,764,946 | 410,850 | 99,160 | 379,570 | 1,183,511 | 2,192,239 | 325,750 |
| Ghana | 2012 | 5.44 | 772,168 | 4,216,564 | 1,358,801 | 2,223,654 | 772,168 | - | - | - | - |
| Ghana | 2015 | 5.55 | 1,214,137 | 6,630,016 | 5,222,663 | 3,496,417 | 1,214,137 | - | - | - | - |
| Ghana | 2016 | 5.22 | 215,026 | 667,868 | 1,024,870 | 352,208 | 215,026 | - | - | - | - |
| Ghana | 2018 | 5.59 | 4,645,570 | 17,724,492 | 10,740,092 | 9,347,219 | - | 4,645,570 | 17,724,492 | 10,740,092 | 9,347,219 |
| Guinea | 2013 | 1.74 | 612,953 | 2,338,329 | 991,322 | 3,192,053 | - | 612,953 | 2,338,329 | 991,322 | 3,192,053 |
| Guinea | 2016 | 1.97 | 1,287,908 | 4,514,146 | 2,281,778 | 2,694,430 | - | 1,287,908 | 4,514,146 | 2,281,778 | 2,694,430 |
| Guinea | 2019 | 2.27 | 1,310,230 | 4,592,385 | 3,574,198 | 3,286,694 | - | 1,310,230 | 4,592,385 | 3,574,198 | 3,286,694 |
| Kenya | 2015 | 6.43 | 1,336,155 | 6,070,536 | 10,234,303 | 2,505,670 | 1,336,155 | - | - | - | - |
| Kenya | 2017 | 6.86 | 2,328,694 | 7,377,031 | 13,043,818 | 3,599,862 | 2,328,694 | - | - | - | - |
| Liberia | 2015 | 1.49 | 816,743 | 3,989,649 | 4,140,424 | 4,050,891 | - | 816,743 | 3,989,649 | 4,140,424 | 4,050,891 |
| Liberia | 2018 | 1.58 | 522,220 | 1,318,871 | 1,120,144 | 897,977 | - | 522,220 | 1,318,871 | 1,120,144 | 897,977 |
| Malawi* | 2012 | 1.47 | 273,377 | 1,073,147 | 824,431 | 366,384 | 273,377 | - | - | - | - |
| Malawi* | 2016 | 1.51 | 865,351 | 3,331,716 | 5,930,353 | 932,364 | - | 865,351 | 3,331,716 | 5,930,353 | 932,364 |
| Malawi* | 2018 | 1.53 | 1,358,563 | 4,341,343 | 6,353,688 | 1,467,507 | - | 1,358,563 | 4,341,343 | 6,353,688 | 1,467,507 |
| Mozambique | 2017 | 6.12 | 1,675,100 | 4,422,101 | 7,842,742 | 2,081,042 | - | 1,675,100 | 4,422,101 | 7,842,742 | 2,081,042 |
| Mozambique | 2020 | 6.37 | 1,882,498 | 4,814,344 | 10,194,846 | 4,531,264 | - | 1,882,498 | 4,814,344 | 10,194,846 | 4,531,264 |
| Niger | 2015 | 2.29 | 38,606 | 203,832 | 29,837 | 305,225 | 38,606 | - | - | - | - |
| Rwanda* | 2013 | 0.89 | 291,991 | 1,594,470 | 1,526,195 | 316,494 | - | 291,991 | 1,594,470 | 1,526,195 | 316,494 |
| Rwanda* | 2016 | 0.87 | 137,104 | 700,088 | 940,959 | 171,909 | 137,104 | - | - | - | - |
| Rwanda* | 2017 | 0.87 | 146,836 | 697,739 | 765,871 | 208,825 | 146,836 | - | - | - | - |
| Rwanda* | 2020 | 0.86 | 870,998 | 2,621,730 | 3,384,902 | 948,856 | 813,106 | 57,892 | 174,257 | 224,981 | 63,066 |
| Senegal | 2013 | 2.73 | 64,388 | 230,423 | 262,239 | 66,336 | 64,388 | - | - | - | - |
| Senegal | 2014 | 2.72 | 255,257 | 913,479 | 1,100,436 | 251,027 | 255,257 | - | - | - | - |
| Senegal | 2016 | 2.69 | 2,249,195 | 8,049,115 | 11,361,351 | 2,027,598 | - | 2,249,195 | 8,049,115 | 11,361,351 | 2,027,598 |
| Senegal | 2019 | 2.59 | 2,118,146 | 5,833,170 | 9,486,467 | 3,126,371 | - | 2,118,146 | 5,833,170 | 9,486,467 | 3,126,371 |
| Sierra-Leone* | 2017 | 1.71 | 782,010 | 2,473,438 | 1,255,591 | 1,763,867 | - | 782,010 | 2,473,438 | 1,255,591 | 1,763,867 |
| Sierra-Leone* | 2020 | 1.73 | 2,615,175 | 9,528,030 | 11,305,371 | 9,912,148 | - | 2,615,175 | 9,528,030 | 11,305,371 | 9,912,148 |
| United Republic of Tanzania | 2015 | 8.10 | 2,605,123 | 10,811,569 | 21,070,784 | 11,304,896 | 2,605,123 | - | - | - | - |
| United Republic of Tanzania | 2020 | 8.61 | 2,255,572 | 7,161,018 | 6,720,358 | 2,814,953 | 2,255,572 | - | - | - | - |
| Togo* | 2011 | 1.29 | 301,546 | 1,370,324 | 880,630 | 1,307,348 | 301,546 | - | - | - | - |
| Togo* | 2014 | 1.32 | 745,100 | 3,220,610 | 1,862,738 | 1,615,507 | - | 745,100 | 3,220,610 | 1,862,738 | 1,615,507 |
| Togo* | 2017 | 1.34 | 633,983 | 2,288,092 | 1,909,034 | 1,463,599 | - | 633,983 | 2,288,092 | 1,909,034 | 1,463,599 |
| Uganda* | 2013 | 4.74 | 194,805 | 616,161 | 382,826 | 84,430 | 194,805 | - | - | - | - |
| Uganda* | 2017 | 5.54 | 3,191,946 | 9,707,244 | 11,507,549 | 2,444,194 | - | 3,191,946 | 9,707,244 | 11,507,549 | 2,444,194 |
| Uganda* | 2020 | 6.16 | 2,648,366 | 8,957,896 | 15,628,773 | 4,906,907 | - | 2,648,366 | 8,957,896 | 15,628,773 | 4,906,907 |
| Zambia | 2014 | 3.37 | 791,488 | 3,036,568 | 4,246,616 | 1,165,462 | 622,580 | 168,908 | 648,021 | 906,251 | 248,716 |
| Zambia | 2017 | 3.67 | 2,423,412 | 7,293,232 | 12,946,583 | 3,118,692 | - | 2,423,412 | 7,293,232 | 12,946,583 | 3,118,692 |
| Zambia | 2020 | 3.96 | 254,302 | 900,186 | 865,123 | 542,875 | 254,302 | - | - | - | - |

* Unknown proportion of funds spent on RDTs vs. ACTs in 2021 from World Malaria Report 2022

**Table S3.** Variables used in the modern housing definition.

| **Variable** | **Type** | **Details of Measurement** |
| --- | --- | --- |
| Improved Floor | Categorical (yes/no) | Improved floor is categorized as having a finished floor (i.e., parquet or polished word, vinyl, ceramic tiles, cement, or carpet) |
| Improved Wall | Categorical (yes/no) | Improved wall is categorized as having a finished wall (cement, stone, bricks, covered adobe, tile, or concrete) |
| Improved Roof | Categorical (yes/no) | Improved roof is categorized as having a finished roof (metal, calamine/cement fiber, ceramic tiles, cement, or asbestos) |
| Modern House | Categorical (yes/no) | Compositive variable of having an improved floor, roof, and walls. |

**Table S4.** Environmental data sources.

| **Environmental variables** | **Details of measurement** | **Resolution** | **Source** |
| --- | --- | --- | --- |
| Total precipitation | Spatial domain: Africa. Monthly.  Cumulative monthly precipitation (mm) | 0.05° x 0.05°  (aprox. 5.6 km) | Climate Hazards Group (CHIRPS)^16^  <https://pubs.usgs.gov/ds/832/> |
| Enhanced vegetation index (EVI) | A measure of vegetation greenness on a scale of 0 to 1 (most green); monthly average, lagged by two months | 5km x 5km | MAP gap-filled EVI  [Oxford MAP EVI: Malaria Atlas Project Gap-Filled Enhanced Vegetation Index  \|  Earth Engine Data Catalog  \|  Google for Developers](https://developers.google.com/earth-engine/datasets/catalog/Oxford_MAP_EVI_5km_Monthly) |
| Average relative humidity at 2m | This parameter is the water vapor pressure as a percentage of the value at which the air becomes saturated (the point at which water vapor begins to condense into liquid water). For temperatures over 0°C (273.15 K) it is calculated for saturation over water. Between -23°C and 0°C this parameter is calculated by interpolating between the ice and water values using a quadratic function. | 0.25° x 0.25°  (aprox. 27km) | ECMWF ERA5^17^  [Monthly averaged data on pressure levels](https://cds.climate.copernicus.eu/cdsapp#!/dataset/reanalysis-era5-pressure-levels-monthly-means?tab=form) |
